# Priming effects of ketone monoester supplementation on theta-burst stimulation-induced plasticity in the primary motor cortex: a randomized placebo-controlled crossover study

**DOI:** 10.64898/2026.09.09.26362322

**Authors:** Abdulhameed Tomeh, Abdul Hanif Khan Yusof Khan, Zalina Abu Zaid, King-Hwa Ling, Liyana Najwa Inche Mat, Hamidon Basri, Muhammad Husni Abu Bakar, Nurul Iman Sofea Suhaimi, Wan Aliaa Wan Sulaiman

## Abstract

Neurological and cardiovascular disorders frequently coexist, yet targeted dual-system interventions remain underinvestigated. Transcranial magnetic stimulation (TMS), including intermittent and continuous theta-burst stimulation (iTBS/cTBS), non-invasively modulates primary motor cortex (M1) excitability. Concurrently, ketone monoesters (KME) show neuroprotective potential and alter cardiovascular hemodynamics. In a randomized, double-blind, placebo-controlled crossover study, 44 healthy young adults were stratified to receive iTBS or cTBS (n = 22/group). Participants completed two sessions separated by 1 week, consuming either a KME or placebo beverage, followed by TBS 1-hour post-ingestion. Motor evoked potentials (MEPs) from the right first dorsal interosseous muscle were tracked for 30 minutes post-stimulation. Blood pressure was measured every 3–5 minutes, heart rate was monitored continuously, and venous blood was sampled for glucose, β-hydroxybutyrate (BHB), pro-BDNF, mature BDNF (mBDNF), and the BDNF rs6265 polymorphism. We found that KME significantly elevated blood BHB and heart rate while decreasing blood glucose and diastolic blood pressure. KME did not alter resting motor threshold, baseline MEP amplitude, or MEP latency. Factorial analysis showed no significant modification of post-TBS MEP time courses by KME. In exploratory within-condition analyses, placebo-cTBS induced significant late increases in MEP amplitude, whereas KME-cTBS showed a blunted post-stimulation response. Serum mBDNF and pro-BDNF concentrations remained unchanged. In conclusion, combining KME and TBS is feasible, well-tolerated, and may exert concurrent neurological and cardiovascular effects. This work opens the door to a novel metabolic-neuromodulatory avenue that merits confirmation in larger cohorts and establishment of clinical relevance in patients with comorbid neurological and cardiovascular disorders.

## 1. Introduction

Nutritional ketosis is a century-old regimen used primarily to treat pediatric epilepsy (Wheless, 2008). Traditionally, ketosis was achieved through a ketogenic diet (KD), consisting of high-fat, low-carbohydrate, and adequate-protein intake, to induce ketone body production by the liver (Poff et al., 2020). The resultant ketone bodies, predominantly β-hydroxybutyrate (BHB), can be utilized as a preferable source of energy substrate by the brain (Hasselbalch et al., 1996; Kapogiannis & Avgerinos, 2020; Mikkelsen et al., 2015). Additionally, the ketone body BHB can alter the dynamics of neurotransmitters to favour higher GABA/glutamate ratio and reduce the occurrence of seizures in epileptic patients (Pflanz et al., 2019; Qiao et al., 2024). On the other hand, the ketone body BHB is increasingly shown as a signaling metabolite that modifies the epigenetic expression of various proteins (Newman & Verdin, 2017). Among these proteins, brain-derived neurotrophic factor (BDNF) has been of particular interest given its pleiotropic roles in neuronal survival and synaptic transmission and plasticity (Kowiański et al., 2018; Y. Li et al., 2022). Pre-clinical studies, both in-vitro and in-vivo, have unanimously shown that the ketone body BHB upregulates the BDNF genetic expression via distinct molecular pathways (Giacco et al., 2026; Hu et al., 2020; Hu et al., 2018; Kwak et al., 2021; Marosi et al., 2016; Sleiman et al., 2016; Sun et al., 2022; Trotta et al., 2022; Zhang et al., 2022). Given these promising neuroprotective effects, recent bibliometric analyses reveal that employing ketogenic diets as treatment is continuously expanding well beyond epilepsy to involve various neurological diseases (Liao et al., 2026; Wang et al., 2022; Yan et al., 2025). However, the frequent reports of “Keto-Flu” (Bostock et al., 2020), along with the strict cut-off for carbohydrate intake represent major issues with compliance to ketogenic diets (Poff et al., 2020). Therefore, scientists aimed to mimic the neuroprotective benefits of ketogenic diets and circumvent their adverse effects through a pharmaceutical product (Rho & Sankar, 2008).

Ketone monoester (KME) ready-to-drink beverage was first investigated in humans in the study of Clarke et al. (Clarke et al., 2012). In their study, KME administration elevated blood ketone BHB acutely within 20 minutes and returned to baseline after ≈ 4 hours of ingestion. The KME ingestion generates a mild state of metabolic ketosis where BHB levels are < 5 mM/L. This is unlike severe metabolic ketosis, e.g., diabetic ketoacidosis, where BHB levels accumulate to more than 20 mM/L due to pathological insulin resistance, followed by a life-threatening decline in blood pH (Kolb et al., 2021). Currently, the KME is mainly sold as an energy drink for athletes after it became certified by the Food and Drug Administration (FDA) in 2015 with GRAS status (Generally Recognized as Safe) (Miyatsu et al., 2024).

The BHB elevation following KME ingestion was shown to serve as a preferential source of energy for exercising muscles (Cox et al., 2016). In addition, the KME-induced ketosis yielded favorable hemodynamic effects, manifesting in an increased heart rate and cardiac output (Oneglia et al., 2023; Selvaraj et al., 2022). Neurologically, the pro-cognitive effects of KME have been frequently reported in athletes with induced mental fatigue (Coleman et al., 2021; Evans & Egan, 2018; Quinones & Lemon, 2022), in adults with obesity (Walsh et al., 2021), and is being currently investigated in elderly populations (Avgerinos et al., 2022). Furthermore, a single dose of KME was reported to enhance the endurance exercise performance in patients with Parkinson’s disease at Hoehn and Yahr stages 1–2 (Norwitz et al., 2020). However, despite the promising neuroprotective potential of the KME supplement, there are no available neurophysiological studies to date to verify its effect on the brain objectively through validated neurophysiological techniques. Throughout recent years, transcranial magnetic stimulation (TMS) has been increasingly used to assess and modulate cortical excitability non-invasively through the scalp (Rossini et al., 2015; Siebner et al., 2022). Among cortical regions, the primary motor cortex (M1) has been of particular interest given the clear behavioural response following its stimulation, represented by muscle twitching that can be recorded via surface electromyography (EMG) (Rossini et al., 2015). As such, TMS-EMG has been an invaluable tool to assess the impact of various pharmacological interventions on the human brain (Sohn et al., 2024; Ziemann et al., 2015). Among the TMS protocols, single- and paired-pulse TMS protocols are used to probe corticospinal and intracortical excitability, respectively. Repetitive TMS (rTMS) protocols are used to induce cortical plasticity, resulting in enhanced or suppressed cortical excitation beyond the stimulation period (Rossini et al., 2015). Theta-burst stimulation (TBS) is a novel and more time-efficient modality of the rTMS with comparable efficacy (Huang et al., 2005). Intermittent (iTBS) and continuous (cTBS) protocols were shown to enhance or suppress M1 excitability, respectively, for several minutes post-stimulation in comparison to baseline (Chung et al., 2016; Huang et al., 2005). At the cellular level, these effects are thought to depend on principles of long-term potentiation/long-term depression (LTP/LTD)- synaptic plasticity (Huang et al., 2017), which in turn rely on the activity-dependent secretion of mature BDNF/pro-BDNF in the synapses, aka the ‘yin and yang’ BDNF hypothesis (Brunoni et al., 2008; C. W. Lee et al., 2023; Lu et al., 2005). In addition to BDNF, the glutamatergic and GABAergic neurotransmissions also play a vital role in the mechanism of iTBS and cTBS (Li et al., 2019), yet at various levels according to the brain region (Stoby et al., 2022). On the other hand, given the intrinsic relationship between the brain and the heart, a growing body of evidence suggests a potential to modulate the cardiovascular functions through delivering TMS to the brain (H. Lee et al., 2023; Schmausser et al., 2022). In fact, it was once hypothesized that delivering TMS to the brain might be a novel strategy to treat arterial hypertension (Cogiamanian et al., 2010). As such, various brain regions have been targeted with different stimulation protocols and heterogeneous results (H. Lee et al., 2023; Schmausser et al., 2022).

Correspondingly, this study aimed to explore the mechanistic convergence of KME and TBS on the human heart and brain physiology for the first time in the literature. The primary objective was to determine whether acute KME supplementation could modify the magnitude of iTBS- and cTBS-induced MEP responses. We hypothesised that KME would enhance the bidirectional TBS-induced plasticity, putatively through BDNF upregulation. Secondary objectives were to assess the effects of KME on baseline corticospinal excitability, metabolic biomarkers, and cardiovascular responses.

## 2. Materials and methods

### 2.1. Study design and protocol registration

This was a randomized, double-blind, placebo-controlled, partial crossover study in accordance with the CONSORT 2010 extension to crossover designs (Dwan et al., 2019). The study was conducted in the neurophysiology laboratory at Hospital Sultan Abdul Aziz Shah. Biochemical analyses of blood samples were conducted in the medical genetics lab at the Faculty of Medicine and Health Sciences of University Putra Malaysia. Study procedures conformed to the Declaration of Helsinki, and ethical approval was obtained from the local Institutional Review Board, approval number JKEUPM-2021-485. The study protocol was preregistered at ClinicalTrials.gov, ID: NCT06799260, and deidentified raw datasets are freely available on the Open Science Framework repository at https://doi.org/10.17605/OSF.IO/R97CG.

### 2.2. Discussion of study design

This was a partial crossover study. The study protocol involved two interventions administered sequentially during each visit; a supplement intervention (KME/placebo) followed after one hour by a stimulation intervention (iTBS/cTBS). The study involved two separate groups of subjects undergoing different TBS protocols (iTBS vs. cTBS). The subjects in each group crossed over to the supplement intervention (KME and placebo) across two visits separated by at least one week.

### 2.3. Subjects

#### 2.3.1. Sample size

G*power (version 3.1.9.7) software was used to calculate the sample size for a mixed ANOVA: Repeated measures, within-between interaction (Faul et al., 2007). Considering an effect size f = 0.2, an α = 0.05, a power (1 - β) = 0.9, two groups, and six repeated MEP measures, a minimum sample size of 36 was calculated. Consequently, we aimed to recruit 44 participants (22 per group) to account for potential dropouts or unmeasurable data.

#### 2.3.2. Selection criteria

Several confounding factors reported in the literature were considered in the following selection criteria to minimize confounding and improve homogeneity of the sample. Inclusion criteria were healthy young adults (18-35 years old) (Corp et al., 2021; Corp et al., 2020), right-handed according to the Edinburgh Handedness Questionnaire (Oldfield, 1971), and fully vaccinated against COVID-19. Exclusion criteria were subjects with contraindications to TMS based on the screening 13-item questionnaire (Rossi et al., 2011), highly active subjects, defined as performing >150 minutes per day of moderate-to-vigorous aerobic activity on at least 5 days per week (Cirillo et al., 2009; Craig et al., 2003), obese individuals, defined as having a Body Mass Index (BMI) ≥ 30 kg/m2 (Sui et al., 2020), smokers (Lang et al., 2008), and individuals with an active or previous lab-confirmed COVID-19 with long symptoms (Manganotti et al., 2023; Ortelli et al., 2022).

#### 2.3.3. Visits and instructions

The study involved three visits to the lab. Visit 1 was for screening and obtaining written informed consent. Subjects were asked to fill out three short questionnaires: the Edinburgh Handedness Questionnaire, the International Physical Activity Questionnaire, and the 13-Item Questionnaire for TMS Safety. Body height and weight were measured using standardized methods, and BMI was calculated. Subjects were offered a short mock experiment to familiarize themselves with the setting and TMS sensations. Visits 2 and 3 involved two experimental conditions separated by at least one week to avoid any carryover effects (Fried et al., 2017). Experiments during visits 2 and 3 were performed at the same time of day (±2 h) in the afternoon for each participant. This was done to minimize M1 plasticity variation due to diurnal fluctuations in cortisol (Sale et al., 2008) and BDNF (Piccinni et al., 2008). Participants were asked to refrain from strenuous exercise, alcohol and caffeine consumption 24 hours before the testing session, get a night sleep ≥ 6 hr, and fast for ≥ 4 hours to minimize the effect of dissimilar food intake on KME gut absorption (Pellegrini et al., 2020; Stubbs et al., 2017). None of the participants were under medications known to influence cortical excitability and plasticity (Sohn et al., 2024; Ziemann, 2004; Ziemann et al., 2015).

#### 2.3.4. Randomization and blinding

Participants (n =44) were randomized to the sequence of interventions in their both visits with an online random number generator at https://www.randomizer.org/.

A third-party researcher, masked to the study hypothesis and objectives, labeled the KME and placebo drink bottles as A or B. The labeled bottles were identical opaque plastic containers. Unblinding the code of the drink condition occurred once all data analyses were complete.

### 2.4. Supplement preparation and dosage

Participants ingested either 500 mg/kg body weight of KME or taste-matched placebo drink (containing arrowroot extract, malic acid, and denatonium benzoate). We chose the dose of 500 mg/kg of KME as it was expected to increase the blood ketone (BHB) levels to ˃ 3 mmol/L within one hour of ingestion and return to baseline after 4 hours (Stubbs et al., 2017). This blood level is sufficient to deliver BHB to the brain through the blood-brain barrier (Hasselbalch et al., 1996; Mikkelsen et al., 2015). All supplements were formulated with cloudy lemonade (Waitrose) to a total volume of 120 mL to improve palatability (Valenzuela et al., 2020). In addition, participants wore a nose clip while consuming the supplements to further mask their bitterness. Both the KME and placebo drinks were administered in identical opaque plastic containers with identical volumes, labeled A and B by a third-party investigator.

### 2.5. Transcranial magnetic stimulation (TMS)

#### 2.5.1. Single pulse TMS

All sessions were performed in the afternoon between 11:00 am and 4:00 pm. Participants were seated in a comfortable motorized Lemi® chair with headrest and armrests. The head position was fixed using a cheek-rest attachment on the chair. Room temperature was maintained at 18–20°C at all times. EMG activity was recorded from the right FDI muscle using disposable pre-gelled Ag-AgCl surface electrodes (P/N 019-435300, Natus, USA). These electrodes were attached to a reusable alligator-clip lead wire (P/ N 117-401900, Natus, USA) and connected to an 8-channel amplifier (AT2 +6, Natus, USA). Participants were seated without visual access to the TMS and EMG equipment displays. After cleansing the skin with alcohol swabs, the active/negative EMG electrode was placed on the skin overlying the FDI belly, while the reference/positive electrode was placed on the metacarpophalangeal joint of the index finger, and the ground electrode was placed on the ulnar styloid process of the wrist. We asked the participants to relax their FDI muscle during all measurements. Raw EMG signals were amplified (AT2 +6, Natus, USA), band-pass filtered from 1 Hz to 10 kHz, digitized at 48 kHz (Nicolet EDX, Natus, USA), and stored on the laboratory computer for offline analysis (Synergy EDX software, version 22.4.1.123, Natus, USA). All participants were offered earplugs to decrease the noise level generated by the TMS coil, as per safety guidelines. All pulses were delivered as biphasic, i.e., anterior-posterior followed by posterior-anterior (AP-PA) current direction in the coil handle, using a butterfly figure-of-eight Cool-B70 coil connected to a MagPro R30 stimulator with add-on theta-burst option (MagVenture A/S, Farum, Denmark).

First, the right FDI motor hotspot was localized by moving the coil in millimeter deviations around the C3h position (Kim et al., 2023). The TMS intensity was first set at 50% maximum stimulator output (MSO). This intensity is thought to be moderately suprathreshold, i.e., elicit EMG activity from the FDI muscle according to previous studies that measured RMT with biphasic pulses through the Cool-B70 coil (Cole et al., 2021). The “motor hotspot” was defined as the optimal coil position, tilt, and orientation that consistently elicited MEPs of maximum amplitude from the relaxed FDI muscle. Although the majority of coil orientations in our study corresponded to the conventional 45° away from the midsagittal line with the handle pointing backwards and laterally in a clockwise direction, some cases deviated more toward 0° or 90°, consistent with previous observations on biphasic pulses (Balslev et al., 2007). Following motor hotspot localization, the coil holder (Super Flex Arm, MagVenture A/S, Farum, Denmark) was locked to fix the coil in position. In addition, we marked the outer shape of the coil with a felt-tipped marker on a mesh swimming cap worn by the participants. This ensured consistent coil placement throughout the session.

The first TMS measure was the resting motor threshold (RMT) using the MTAT 2.1 software, available on http://www.clinicalresearcher.org/software.htm. At the optimal FDI hotspot, TMS intensity was decreased to 37% MSO. The intensity was then adjusted based on the success or failure in obtaining an EMG response ≥ 50 microvolts (μV). Thirty single pulses were applied to measure the RMT as recommended (Borckardt, 2022; Koponen & Peterchev, 2022). The RMT was measured twice in each session: immediately prior to supplementation (RMT_1_) and at 1-hour post-supplementation (RMT_2_), when blood BHB concentrations were expected to peak.

The second TMS measure was the MEP amplitude in μV, operationally defined as the voltage difference between peak-to-trough (P. Li et al., 2022), i.e., negative to positive deflections of the MEP wave, commonly referred to as peak-to-peak amplitude (Rossini et al., 2015). And the third measure was the MEP latency in milliseconds, defined as the time elapsed between the TMS artifact and the beginning of the MEP trace (Milardovich et al., 2023). In each MEP measurement, we calculated the average of 12 MEPs elicited at 120% RMT intensity and jittered between 5 and 15 seconds to minimize anticipation by the participants, and consequently avoid voluntary muscle contraction (Capozio et al., 2021; Tran et al., 2021). A successful MEP block was defined as one where at least six automatically measurable MEP recordings were obtained by the EMG machine out of the twelve stimuli. This was based on evidence from two meta-analyses converging on the notion that > 5 stimuli are required to produce an excellent within-session MEP amplitude reliability (Cavaleri et al., 2017; Osnabruegge et al., 2023). However, in a minority of cases, if six automatically measurable MEP recordings were not obtained due to subject head movement or the temporal dispersion and phase cancellation phenomenon (Kimura, 2019), the coil positioning was readjusted, and an additional block of 12 stimuli was obtained. All automatically measurable MEP recordings from the two blocks were averaged and included in the analysis.

We measured the MEP blocks at eight timepoints: pre-supplementation (T_pre-sup_), at 1-hour post-supplementation (T_post-sup_), pre-TBS (T_0_), and post-TBS at 0-3 (T_1_), 4-7 (T_2_), 9-12 (T_3_), 17-20 (T_4_), and 27-30 minutes (T_5_). The TMS intensity used to collect MEP blocks at T_pre-sup_ and T_post-sup_ was set at 120% RMT_1_, whereas the TMS intensity to collect MEP blocks at T_0_, T_1_, T_2_, T_3_, T_4_, and T_5_ was set at 120% RMT_2_. In each timepoint post-TBS, the mean value of MEPs was averaged and compared to pre-TBS (T_0_) using the following equation: (conditioned MEP amplitude/T_0_ MEP amplitude) ×100. A value of 90–110 % represents no change, while values <90 % represent suppression, and >110 % represent facilitation of the corticospinal excitability following TBS (Boucher et al., 2021). Grand average MEP (MEPGA) is defined as the mean value of the %change in MEP amplitude at T_1_, T_2_, T_3_, T_4_, and T_5_. To maintain a relatively fixed level of vigilance, participants viewed episodes from a National Geographic documentary, following the methodology of a previous study (McCalley et al., 2021). To evaluate unintended TMS-related sensations, participants completed the TMSens_Q questionnaire at the end of each session (Giustiniani et al., 2022).

#### 2.5.2. Intermittent and continuous theta-burst stimulation

Two theta-burst stimulation (TBS) protocols were applied, namely intermittent TBS (iTBS) and continuous TBS (cTBS), in two different groups of subjects. The iTBS protocol consisted of a 2-second TBS train of pulses repeated every 10 seconds for 192 seconds, while the cTBS protocol consisted of a continuous TBS train of pulses for 40 seconds. Each TBS train comprised a 3-pulse burst spaced 20 ms apart (i.e., 50 Hz) and was repeated every 200 ms (i.e. 5 Hz), for a total of 600 pulses (Huang et al., 2005). Stimulation intensity was set at 70% RMT_2_ for each subject (Fried et al., 2019; Goldsworthy et al., 2014).

### 2.6. Cardiovascular monitoring

Participants were asked to empty their bladders before the session and rest in the laboratory for 3-5 minutes without talking or moving around (Muntner et al., 2019). Blood pressure (BP) was measured serially every 3-5 minutes using a fully automated, oscillometric sphygmomanometer (G30E, Philips Goldway Industrial Inc., Shenzhen, China) while the participants sat in the TMS chair in a semi-recumbent position. Heart rate (HR) was monitored continuously via a pulse oximeter. The monitor screen was video-recorded for later analysis. For data reduction, HR values were extracted at 30-second intervals (Skinner et al., 2017). To ensure data integrity, HR measurements obtained during automated blood pressure cuff inflation were excluded from the analysis due to potential pressure-induced signal interference (Sondej & Zawadzka, 2022). Readings of systolic BP (SBP), diastolic BP (DBP), mean arterial pressure (MAP), and HR were averaged and grouped into five timepoints: pre-supplementation (T_pre-sup_), at 1 hour post-supplementation (T_post-sup_), the first 10 minutes post-TBS (T_10min_), the second 10 minutes post-TBS (T_20min_), and the third 10 minutes post-TBS (T_30min_).

A schematic diagram of the study protocol is shown in Fig. 1.

**Fig. 1.**
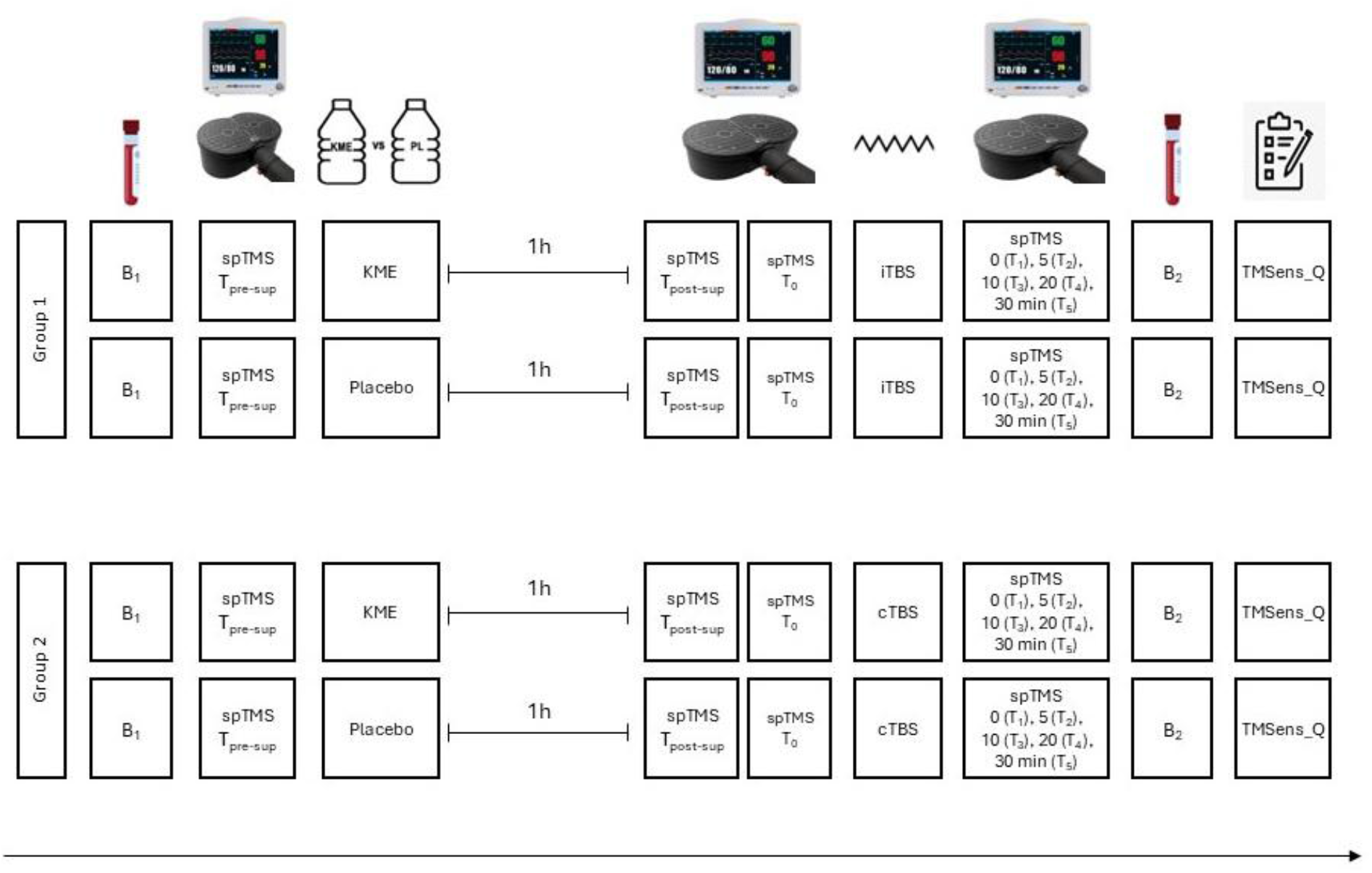
Schematic representation of the study protocol. The study protocol involved two interventions administered sequentially during each visit; supplement intervention (KME/placebo) followed after ⁓ 1h by a stimulation intervention (iTBS/cTBS). There were two separate groups of subjects undergoing different TBS protocols (iTBS vs cTBS). The subjects of each group crossed over to the supplement intervention (KME and placebo) in two visits separated by at least one week. In each visit, measures of corticospinal excitability through single-pulse TMS (spTMS) were obtained before the supplement (T_pre-sup_) and at 1h post-supplementation (T_post-sup_) at 120% of resting motor threshold (RMT_1_) intensity that was measured at T_pre-sup_. Following that, RMT was adjusted after the supplement (RMT_2_), and measures of corticospinal excitability through spTMS were obtained pre-TBS (T_0_), and post-TBS at 0 (T_1_), 5 (T_2_), 10 (T_3_), 20 (T_4_), and 30 minutes (T_5_) at 120% of the adjusted RMT (RMT_2_) intensity that was measured at T_0_. Measures of blood pressure and heart rate were obtained throughout the TMS session, and two blood samples were obtained, at the beginning of session (B_1_) and after the end of all TMS measures (B_2_). TMSens_Q questionnaire was filled out at the end of each session.

### 2.7. Blood chemistry analysis

#### 2.7.1. Blood collection and processing

Two blood samples were collected from the antecubital vein during each visit: one upon arrival and the other at the end of the visit. The first sample of the first visit consisted of 8 mL of blood, which was split into a 5-mL ethylenediaminetetraacetic acid (EDTA) tube and a 3-mL serum separator tube (SST). All subsequent samples consisted of 3 mL of blood collected in SSTs. EDTA samples were stored on ice until the conclusion of the daily TMS sessions, before being transferred to the medical genetics laboratory for DNA extraction. Meanwhile, the SST samples were left to clot at room temperature for 60-120 minutes. This duration ensures that most BDNF is released from platelets into the serum, where it remains relatively stable thereafter (Gejl et al., 2019; Maffioletti et al., 2014). Following the clotting period, SST samples were centrifuged at room temperature for 15 minutes at 1000 × g. Consequently, all samples were aliquoted into duplicates and stored at −80°C until analysis.

#### 2.7.2 Blood glucose and beta-hydroxybutyrate measurement

The whole blood collected in SSTs was used for glucose and BHB measurements. A third-party researcher, who was blinded to the intervention assignments of the participants, measured glucose and BHB on-site using test strips and a portable monitor (FreeStyle Optium Neo, Abbott Laboratories, USA). These values were disclosed to the study researcher at the end of the study.

#### 2.7.3 DNA extraction and BDNF genotyping

EDTA tubes were centrifuged at 4000 rpm for 10 minutes to isolate the buffy coat. Genomic DNA was extracted from the buffy coat using the QIAamp DNA Mini Kit (QIAGEN, Hilden, Germany) according to the manufacturer’s instructions. The quality and concentration of extracted DNA were determined using a NanoVue Plus UV Spectrophotometer (GE Healthcare, Chicago, USA). The DNA samples were then stored at −20°C and analyzed within three to five months.

Primer sequences were designed based on the primer3 database https://primer3.ut.ee/ (Untergasser et al., 2012), as Forward: CGCCGTTACCCACTCACTAATA; and Reverse: CCAGGTGAGAAGAGTGATGACC, producing an amplicon size of 466 bp. Amplification of the BDNF gene was performed via polymerase chain reaction (PCR) using a peqSTAR Thermal Cycler (PEQLAB Biotechnologie GmbH, Germany). Gradient PCR products were electrophoresed on a 2% agarose gel and visualized using a gel documentation system (Syngene GBOX-CHEMI, Thermo Fisher, USA). The final PCR cycling conditions consisted of an initial denaturation for 3 minutes at 95°C, followed by 35 cycles of 95°C for 30 seconds, 61°C for 30 seconds, and 72°C for 60 seconds, and a final extension at 72°C for 5 minutes. PCR products were purified using the PrimeWay Gel Extraction/PCR Purification Kit (1st BASE, Singapore) according to the manufacturer’s instructions. The purified products were sent for Sanger sequencing using services provided by Apical Scientific Laboratories Sdn Bhd, Malaysia. Sequence analysis was performed using Unipro UGENE software, version 52.1 (Okonechnikov et al., 2012), as shown in Fig. 2. Eventually, all participants were successfully genotyped for the rs6265 polymorphism of interest.3

**Fig. 2.**
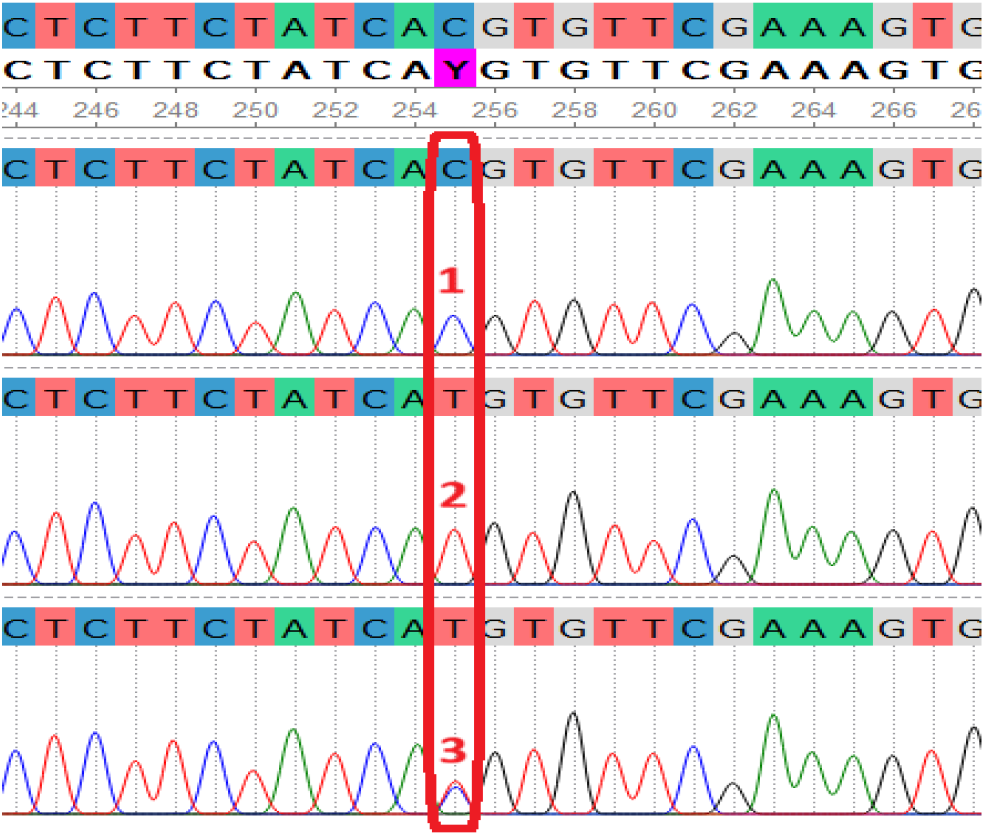
BDNF rs6265 single nucleotide polymorphism. Sanger sequencing chromatograms displaying the nucleotide variations: Subject 1 exhibits a homozygous C/C nucleotide genotype (Val/Val amino acid profile); Subject 2 exhibits a homozygous T/T genotype (Met/Met profile); and Subject 3 displays a heterozygous C/T genotype (Val/Met profile). The top alignment track represents the reference sequence derived from the GRCh38/hg38 Human Genome Assembly at https://genome-asia.ucsc.edu

#### 2.7.4 BDNF serum level quantification

All serum samples were analyzed within 6 months of collection. Although many commercial kits are available for BDNF quantification, several utilize antibodies that cross-react with both mature and pro-BDNF isoforms, leading to heterogeneous results when testing the same blood sample with different ELISA kits. A study evaluating six of the most common BDNF assays found that the Quantikine and Aviscera assays selectively recognized mature BDNF; in contrast, other tested assays reacted with both mature and pro-BDNF, reflecting total BDNF rather than specific isoforms (Polacchini et al., 2015). Accordingly, we used the Quantikine ELISA kit (DBD00, R&D Systems, Minneapolis, USA) for mBDNF and the DuoSet ELISA kit (DY3175, R&D Systems, Minneapolis, USA) for pro-BDNF according to the manufacturers’ instructions. All samples were analyzed in duplicate by the same investigator. Absorbance was measured at 450 nm, with a correction wavelength set at 570 nm, using a VersaMax microplate reader (Molecular Devices, USA). A four-parameter logistic curve was fitted using an online software at https://www.myassays.com/. The optical density values fell outside the standard curve in 55% of pro-BDNF samples (below the curve) and 26% of mBDNF samples (above the curve), and these were consequently excluded from statistical analyses. A picture of pro-BDNF and mBDNF ELISA plates at the end of the assay is shown in Fig. 3.

**Fig. 3.**
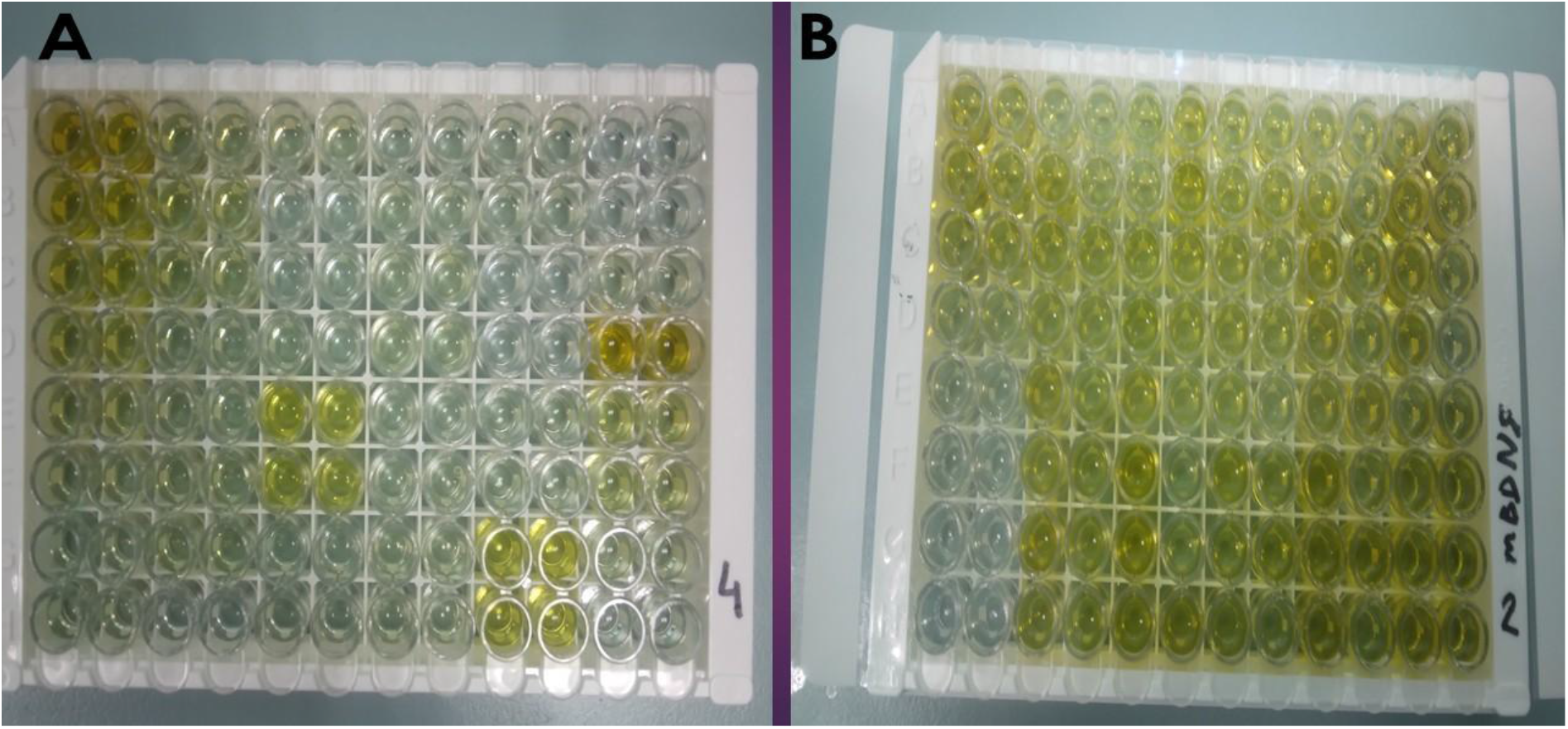
ELISA plates. (A) pro-BDNF and (B) m-BDNF ELISA plates at the end of the assay. Note the low optical density of most pro-BDNF samples (A), indicating that protein concentrations were mostly blow the detection range of the kit.

### 2.8. Data analysis

SPSS version 30 software (IBM Corp., NY, USA) was used for all analyses. Graphs were generated using GraphPad Prism version 10. Data cleaning was initially conducted, which included checking for normality of distribution with the Kolmogorov-Smirnov test and histogram plots, applying proper transformations, removing outliers, and replacing missing values. Cardiovascular (SBP, DBP, MAP, and HR), neurophysiological (RMT and MEP latency), and blood (BHB and glucose) measures were normally distributed (Kolmogorov-Smirnov p-value > 0.05), and no outliers were detected. Serum mBDNF and pro-BDNF values were non-normally distributed with multiple missing values. Therefore, they were analyzed with pairwise, nonparametric tests. MEP amplitudes were not normally distributed and were therefore natural-log transformed, Ln(nMEP amplitude), following the approach of previous studies (Halawa et al., 2021; Hinder et al., 2014; Perellón-Alfonso et al., 2018; Puri et al., 2016). Due to technical issues, four MEP blocks were missing. To account for this, the mean substitution method was used (Gomila & Clark, 2022). As such, the average of the MEP blocks at an individual level was calculated and used to replace the missing values as appropriate.

A three-way mixed repeated-measures analysis of variance (3-way mixed rmANOVA) was used to analyze Ln(nMEP amplitude) and MEP latency separately with factors: TBS (2 levels: iTBS, cTBS) as between-subject factor and TIME (6 levels: T_0_, T_1_, T_2_, T_3_, T_4_, T_5_) and SUPPLEMENT (2 levels: KME, placebo) as within-subject factors to examine the effect of KME on iTBS and cTBS after-effects in comparison to placebo. To rule out any transient, timepoint-specific effects of the KME supplement on raw MEP amplitudes, nonparametric pairwise comparisons were applied at each individual interval and compared to baseline. To examine the effect of BDNF genotype status on Ln(nMEP amplitude), two 3-way mixed ANOVAs were run in the iTBS and cTBS groups separately with factors: GENOTYPE (3 levels: Val/Val, Val/Met, Met/Met) as between-subject factor and TIME (6 levels: T_0_, T_1_, T_2_, T_3_, T_4_, T_5_) and SUPPLEMENT (2 levels: KME, placebo) as within-subject factors to examine the effect of BDNF rs6265 polymorphism on iTBS and cTBS after-effects separately, and whether these effects were further modulated by the KME supplement. A 2×2 rmANOVA with TIME (2 levels: T_pre-sup_, T_post-sup_) and SUPPLEMENT (2 levels: KME, placebo) as within-subject factors was performed to examine the effect of KME on RMT, MEP amplitude, and MEP latency after 1h in comparison to placebo. Separate 3-way mixed rmANOVA tests were run with factors: TBS (2 levels: iTBS, cTBS) as between-subject factor and TIME (5 levels: T_pre-sup_, T_post-sup_, T_10min_, T_20min_, T_30min_) and SUPPLEMENT (2 levels: KME, placebo) as within-subject factors to examine the effect of KME on SBP, DBP, MAP, and HR in comparison to placebo, and whether these effects were further modulated by the TBS protocol. A 3-way mixed ANOVA was run with factors TBS (2 levels: iTBS, cTBS) as between-subject factor and TIME (2 levels: B_1_, B_2_) and SUPPLEMENT (2 levels: KME, placebo) as within-subject factors to examine the effect of KME on blood glucose and BHB, and whether these effects were further modulated based on the TBS protocol. The Greenhouse-Geisser correction was used when the sphericity assumption was violated, as indicated by a significant Mauchly’s test (*p* < 0.05). In case of significant *F*-test results of the interactions, rmANOVA was followed by post hoc analysis using Bonferroni adjustment. Nonparametric Wilcoxon signed-rank tests were used to analyze the differences in serum mBDNF and pro-BDNF from pre- to post-intervention within groups. To explore potential associations between continuous variables, Pearson correlation coefficient (r) was used for parametric data and Spearman correlation (rho) was used for non-parametric data. Normally distributed data are represented as mean ±standard deviation, and non-normally distributed data are represented as median (interquartile range). All *p*-values are two-tailed and considered statistically significant when *p* ≤ 0.05.

## 3. Results

### 3.1. Demographic data of participants

Forty-five healthy young adults were enrolled in this study. One female subject was withdrawn at the beginning of session 1 due to high RMT and safety risks associated with the high-frequency TBS protocol, and her data were consequently excluded from all analyses. Demographic data of the participants are shown in Table 1. The CONSORT flow diagram for subject recruitment is illustrated in Fig. 4.

**Table 1.** Demographic data of participants.

| | Mean ± SD | | |
| --- | --- | --- | --- |
|  | All | iTBS group | cTBS group |
| N | 44 | 22 | 22 |
| Age (years) | 22.1 ± 2.7 | 22.4 ± 3.7 | 21.7 ± 1.1 |
| Gender, F/M | 23/21 | 14/8 | 9/13 |
| Ethnicity, Malay/Chinese/Indian | 32/6/6 | 17/3/2 | 15/3/4 |
| Edinburgh laterality quotient | 76.1 ± 21.1 | 71.8 ± 22.2 | 80.5 ± 19.4 |
| Height (meters) | 164.5 ± 10.3 | 163.1 ± 9.1 | 165.9 ± 11.5 |
| Weight (kg) | 60.1 ± 12.8 | 60.1 ± 14.8 | 60.1 ± 10.9 |
| BMI (kg/m<sup>2</sup>) | 22.1 ± 3.4 | 22.3 ± 3.8 | 21.8 ± 3.1 |
| Vigorous activity days/week | 1.5 ± 1.6 | 1.3 ± 1.4 | 1.7 ± 1.8 |
| Vigorous activity hours/day | 0.7 ± 0.8 | 0.8 ± 0.9 | 0.6 ± 0.7 |
| Moderate activity days/week | 1.7 ± 1.9 | 1.7 ± 1.7 | 1.8 ± 2.2 |
| Moderate activity hours/day | 0.5 ± 0.5 | 0.5 ± 0.5 | 0.5 ± 0.6 |
| Walking 10 min days/week | 6.3 ± 1.6 | 6.1 ± 1.9 | 6.4 ± 1.3 |
| Walking hours/day | 3.6 ± 2.9 | 3.5 ± 2.8 | 3.7 ± 3.1 |
| Sitting hours/day | 6.5 ± 1.9 | 7.4 ± 1.7 | 5.4 ± 1.4 |
| BDNF rs6265 genotype |  |  |  |
| Val-Val/ Val-Met/ Met-Met | 16/20/8 | 6/12/4 | 10/8/4 |

N: number of participants, F: female, M: male, BMI: Body Mass Index, BDNF: Brain-derived neurotrophic factor

**Fig. 4.**
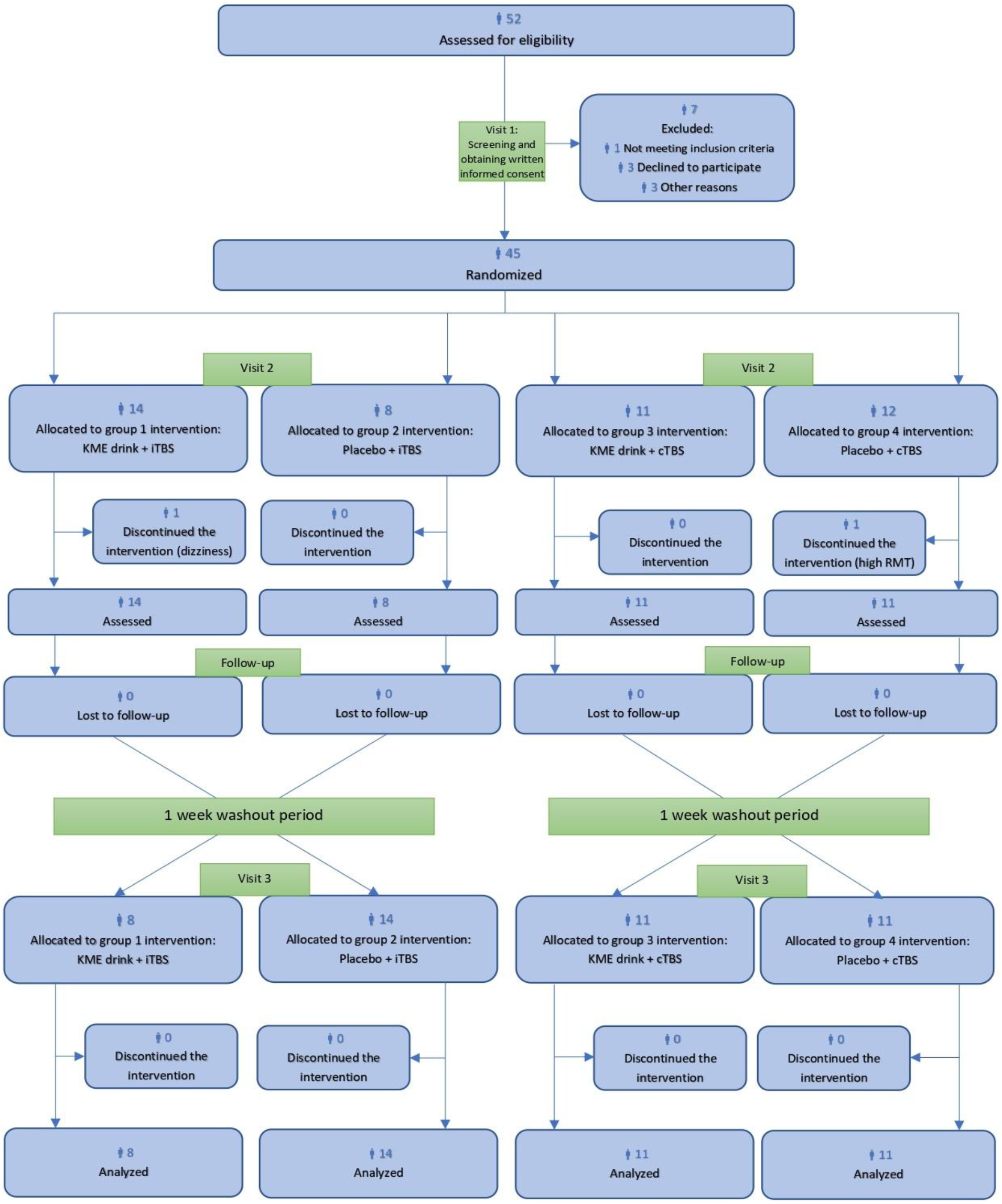
CONSORT flow diagram of participant recruitment.

### 3.2. TMSens_Q Questionnaire

The TMS was well-tolerated across all sessions. One female subject in the KME-iTBS group discontinued the session at 20 minutes post-iTBS (T_4_) due to a complaint of dizziness. This event was associated with a drop in blood pressure to 68/39, compared to a baseline of 108/70. No loss of consciousness occurred. The subject quickly recovered after discontinuing the TMS pulses and acknowledged skipping breakfast and feeling that the TMS pulses over her scalp were noxious. Cases of vasovagal syncope in healthy adults have been documented in the TMS literature (Gillick et al., 2015). The subject was contacted the day after the session and confirmed the absence of any residual symptoms. Overall, sleepiness was the most reported TMSens_Q sensation in 37 subjects (84.1%) and 36 subjects (81.8%) in the KME and placebo groups, respectively. Out of the 88 sessions, eight subjects correctly identified KME and five subjects correctly identified placebo. The questionnaire results are shown in Fig. 5. All sensations were experienced during the TMS delivery and resolved at the end of each session, with the exception of nausea in the KME group, which was reported to persist for 2-3 hours after the session.

**Fig. 5.**
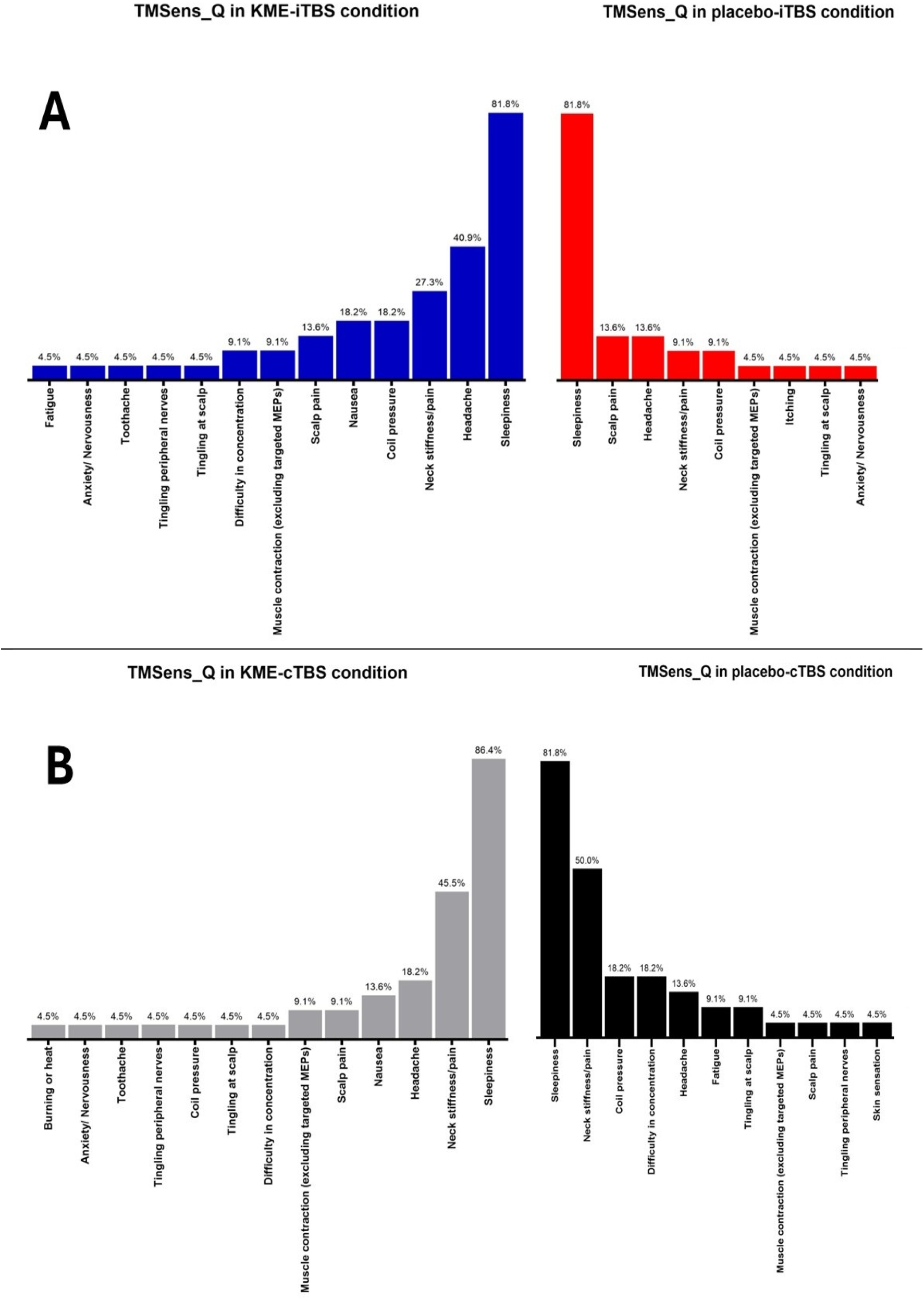
Results of the structured TMSens_Q questionnaire. The graph depicts the frequency of sensations in the iTBS (A) and cTBS (B) groups as demonstrated by the percentage of subjects reporting their sensations at the end of each session.

### 3.3. Effects of KME on baseline corticospinal excitability indices

The 2×2 rmANOVAs of baseline corticospinal excitability indices—including RMT, MEP amplitude, and MEP latency—did not reveal any significant effect of TIME, SUPPLEMENT, or TIME × SUPPLEMENT, p > 0.05. This indicates that the KME supplement did not significantly modulate the baseline corticospinal excitability compared to placebo. Absolute values and full ANOVA results are shown in Table 2 and Table 3, respectively.

**Table 2.** Absolute values of baseline corticospinal excitability indices.

|  | KME group<br>(N = 44) |  | Placebo group<br>(N = 44) |  |
| --- | --- | --- | --- | --- |
|  | T<sub>pre-sup</sub> | T<sub>post-sup</sub> | T<sub>pre-sup</sub> | T<sub>post-sup</sub> |
| RMT (%MSO) | 42.3 ± 7.8 | 42.8 ± 9.1 | 41.8 ± 8.4 | 41.9 ± 8.6 |
| MEP amplitude (μV) | 1136.7 ± 450.3 | 1225.9 ± 961.7 | 1261.9 ± 760.7 | 1234.6 ± 871.1 |
| MEP latency (ms) | 22.16 ± 1.45 | 22.21 ± 1.33 | 22.25 ± 1.57 | 22.31 ± 1.67 |

Values are means ± standard deviation. RMT: resting motor threshold, MEP: motor evoked potential, MSO: maximum stimulator output, μV: microvolts, ms: milliseconds, KME: ketone monoester.

**Table 3.** ANOVA results of baseline corticospinal excitability indices.

|  | TIME | SUPPLEMENT | TIME × SUPPLEMENT |
| --- | --- | --- | --- |
| RMT | $F(1,43) = 0.930$,<br>$p = 0.34$,<br>$\eta_p^2 = 0.021$ | $F(1,43) = 0.935$,<br>$p = 0.339$,<br>$\eta_p^2 = 0.021$ | $F(1,43) = 0.5$,<br>$p = 0.483$,<br>$\eta_p^2 = 0.012$ |
| MEP amplitude | $F(1,43) = 0.149$,<br>$p = 0.701$,<br>$\eta_p^2 = 0.003$ | $F(1,43) = 0.339$,<br>$p = 0.546$,<br>$\eta_p^2 = 0.008$ | $F(1,43) = 0.645$,<br>$p = 0.426$,<br>$\eta_p^2 = 0.015$ |
| MEP latency | $F(1,43) = 0.417$,<br>$p = 0.522$,<br>$\eta_p^2 = 0.01$ | $F(1,43) = 0.954$,<br>$p = 0.334$,<br>$\eta_p^2 = 0.022$ | $F(1,43) = 0.012$,<br>$p = 0.912$,<br>$\eta_p^2 = 0.001$ |

2 × 2 rmANOVAs were performed on RMT, MEP amplitude, and MEP latency, using within-subject factors of SUPPLEMENT (2 levels: KME, placebo) and TIME (2 levels: T<sub>pre-sup</sub>, T<sub>post-sup</sub>). RMT: resting motor threshold, MEP: motor evoked potential, MSO: maximum stimulator output, KME: ketone monoester.

### 3.4. Effects of KME on TBS-induced neuroplasticity

A three-way rmANOVA of Ln(nMEP amplitude) showed a significant effect of TIME [*F* (4.108,172.54) = 5.418, *p* < 0.001, *η_p_^2^* = 0.114] and TIME × TBS [*F* (4.108,172.54) = 3.113, *p* = 0.01, *η_p_^2^* = 0.069]. Bonferroni adjusted pairwise comparisons for the interaction effect of TIME × TBS showed that Ln(nMEP amplitude) in the iTBS group significantly increased at T_1_ in comparison to T_0_ (mean difference = 0.122; 95% CI, 0.003–0.24; *p* = 0.039), and at T_3_ in comparison to T_0_ (mean difference = 0.245; 95% CI, 0.035–0.455; *p* = 0.011); while in the cTBS group, Ln(nMEP amplitude) significantly increased at T_4_ in comparison to T_0_ (mean difference = 0.171; 95% CI, 0.010–0.331; *p* = 0.029), and at T_5_ in comparison to T_0_ (mean difference = 0.220; 95% CI, 0.019–0.420; *p* = 0.022); furthermore, Ln(nMEP amplitude) was significantly higher at T_2_ post-iTBS in comparison to T_2_ post-cTBS (mean difference = 0.163; 95% CI, 0.017–0.309; *p* = 0.03).

To rule out any transient, timepoint-specific effects of KME on MEP amplitudes, pairwise comparisons were applied at each individual interval and compared to baseline. Remarkably, the KME-cTBS condition was the only condition with no significant modulation of MEP amplitude across any timepoint, all *p* > 0.2 (Table 5).

Separate three-way rmANOVAs of Ln(nMEP amplitude) in the iTBS and cTBS groups revealed that the GENOTYPE factor did not have any main or interaction effects, all *p* > 0.05 (Table 7).

Additional three-way rmANOVA showed a significant main effect of TIME only on MEP latency [*F* (3.434,144.23) = 19.74, *p* < 0.001, *η_p_^2^* = 0. 320], but no main or interaction effects of TBS and SUPPLEMENT approached significance, all *p* > 0.05.

Absolute values and full ANOVA results of the TMS data are shown in Table 4 and Table 6, respectively. The time-course changes in corticospinal excitability indices are illustrated in Fig. 6.

**Table 4.** Absolute values of the outcome measures.

| Variables | iTBS group<br>(N = 22) |  | cTBS group<br>(N = 22) |  |
| --- | --- | --- | --- | --- |
|  | KME | Placebo | KME | Placebo |
| MEP amplitude—μV | | | | |
| T<sub>0</sub> | 1229.8<br>(863.4–1778.6) | 1181.9<br>(785.9–1456.9) | 1082.2<br>(800.7–1467.7) | 1034.3<br>(738.0–1558.4) |
| T<sub>1</sub> | 1291.1<br>(836.3–1761.3) | 1325.7<br>(930.7–1799.5) | 981.3<br>(773.1–1716.1) | 1150.2 (779.6–1582.2) |
| T<sub>2</sub> | 1267.1<br>(971.5–1633.4) | 1368.7<br>(792.0–1780.7) | 1019.5<br>(755.9–1598.9) | 938.0 (659.9–1592.1) |
| T<sub>3</sub> | 1553.7<br>(944.9–2620.0) | 1292.2<br>(875.9–2306.6) | 1182.5<br>(786.8–1878.5) | 1153.8<br>(727.1–1993.7) |
| T<sub>4</sub> | 1097.2<br>(901.2–1903.2) | 1155.5<br>(974.1–1591.6) | 1269.0<br>(857.8–1740.1) | 1212.6<br>(888.4–1976.0) |
| T<sub>5</sub> | 1144.7<br>(1023.1–1942.6) | 1253.6<br>(1070.6–1766.4) | 1258.5<br>(910.1–1826.4) | 1471.4<br>(975.7–2072.7) |
| MEP latency—ms |  |  |  |  |
| T<sub>0</sub> | 22.29 ± 1.39 | 22.37 ± 1.53 | 22.48 ± 1.45 | 22.43 ± 1.65 |
| T<sub>1</sub> | 22.39 ± 1.32 | 22.38 ± 1.64 | 22.64 ± 1.65 | 22.55 ± 1.77 |
| T<sub>2</sub> | 22.44 ± 1.48 | 22.49 ± 1.70 | 22.78 ± 1.83 | 22.57 ± 1.80 |
| T<sub>3</sub> | 22.49 ± 1.35 | 22.54 ± 1.75 | 22.91 ± 1.98 | 22.72 ± 1.89 |
| T<sub>4</sub> | 22.76 ± 1.43 | 22.62 ± 1.70 | 22.87 ± 1.67 | 22.81 ± 1.80 |
| T<sub>5</sub> | 22.95 ± 1.50 | 22.92 ± 1.71 | 23.01 ± 1.83 | 22.84 ± 1.84 |

Values are means ± standard deviation for normally distributed data and medians (interquartile range) for non-normally distributed data. iTBS: intermittent theta-burst stimulation, cTBS: continuous theta-burst stimulation, KME: ketone monoester, MEP: motor evoked potential, SBP: systolic blood pressure, DBP: diastolic blood pressure, MAP: mean arterial pressure, HR: heart rate, BHB: beta-Hydroxybutyrate, BDNF: brain-derived neurotrophic factor

**Table 5.** Pairwise comparison of absolute values of TBS-related MEPs.

| Comparisons | iTBS group<br>(N = 22) |  | cTBS group<br>(N = 22) |  |
| --- | --- | --- | --- | --- |
|  | KME | Placebo | KME | Placebo |
| MEP amplitude |  |  |  |  |
| T<sub>1</sub> vs. T<sub>0</sub> | $T = 150, p = 0.445$ | $T = 177, p = 0.101$ | $T = 145, p = 0.548$ | $T = 162, p = 0.249$ |
| T<sub>2</sub> vs. T<sub>0</sub> | $T = 162, p = 0.249$ | $T = 172, p = 0.14$ | $T = 129, p = 0.935$ | $T = 144, p = 0.57$ |
| T<sub>3</sub> vs. T<sub>0</sub> | **$T = 211, p = 0.006$** | $T = 172, p = 0.14$ | $T = 149, p = 0.465$ | $T = 181, p = 0.077$ |
| T<sub>4</sub> vs. T<sub>0</sub> | $T = 163, p = 0.236$ | $T = 176, p = 0.108$ | $T = 165, p = 0.211$ | **$T = 217, p = 0.003$** |
| T<sub>5</sub> vs. T<sub>0</sub> | $T = 185, p = 0.058$ | **$T = 199, p = 0.019$** | $T = 161, p = 0.263$ | **$T = 200, p = 0.017$** |
| MEP latency |  |  |  |  |
| T<sub>1</sub> vs. T<sub>0</sub> | $t = 1.76, p = 0.093$ | $t = 0.176, p = 0.862$ | $t = 1.25, p = 0.225$ | $t = 1.65, p = 0.114$ |
| T<sub>2</sub> vs. T<sub>0</sub> | $t = 1.58, p = 0.129$ | $t = 1.5, p = 0.149$ | $t = 1.892, p = 0.072$ | $t = 1.364, p = 0.187$ |
| T<sub>3</sub> vs. T<sub>0</sub> | $t = 1.698, p = 0.104$ | $t = 1.494, p = 0.15$ | $t = 1.951, p = 0.065$ | **$t = 2.465, p = 0.022$** |
| T<sub>4</sub> vs. T<sub>0</sub> | **$t = 4.214, p < 0.001$** | **$t = 2.203, p = 0.039$** | **$t = 3.43, p = 0.003$** | **$t = 3.786, p < 0.001$** |
| T<sub>5</sub> vs. T<sub>0</sub> | **$t = 5.342, p < 0.001$** | **$t = 5.739, p < 0.001$** | **$t = 3.271, p = 0.004$** | **$t = 3.956, p < 0.001$** |

MEP amplitude at every timepoint was analyzed in comparison to baseline using the non-parametric Wilcoxon matched-pair signed-rank test. MEP latency at every timepoint was analyzed in comparison to baseline using the parametric paired-samples t-test. MEP: motor evoked potential, iTBS: intermittent theta-burst stimulation, cTBS: continuous theta-burst stimulation, KME: ketone monoester. Values in bold denote statistically significant results.

**Fig. 6.**
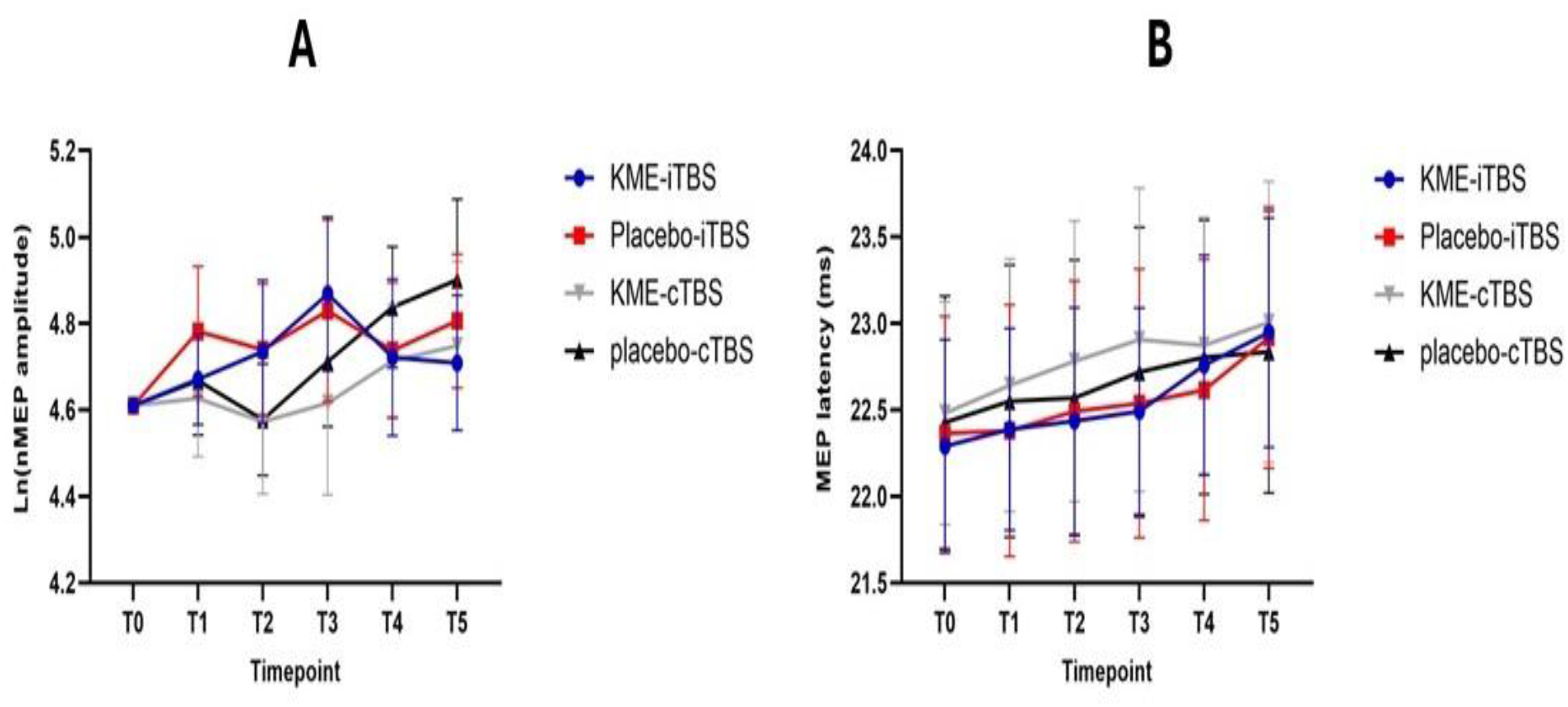
Comparison of corticospinal excitability indices between the different supplement-TBS combinations. Natural log-transformed normalized MEP amplitudes (A) and MEP latencies (B) plotted at every timepoint immediately pre-TBS (T_0_) and post-TBS (T_1_, T_2_, T_3_, T_4_, T_5_). Error bars denote 95 % confidence interval around the mean.

**Table 6.**
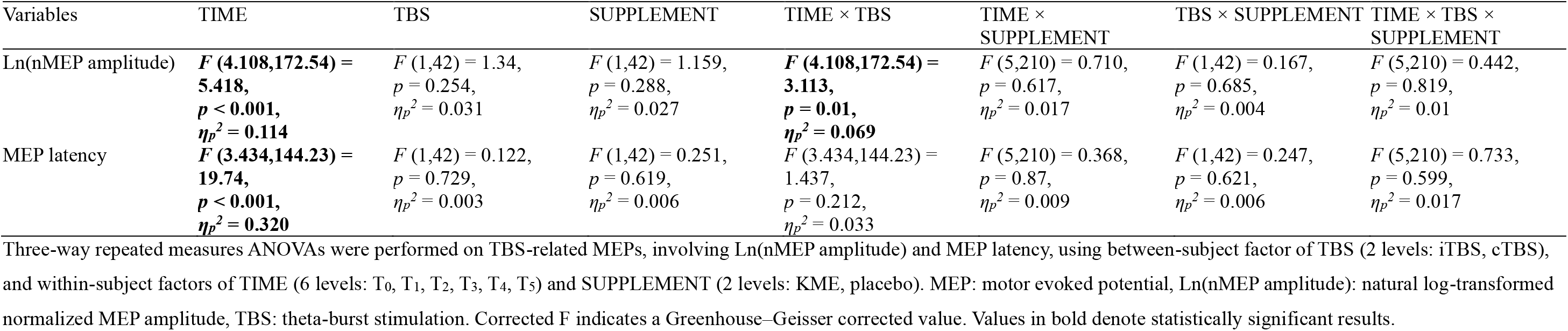
ANOVA results of TBS-related MEPs.

**Table 7.**
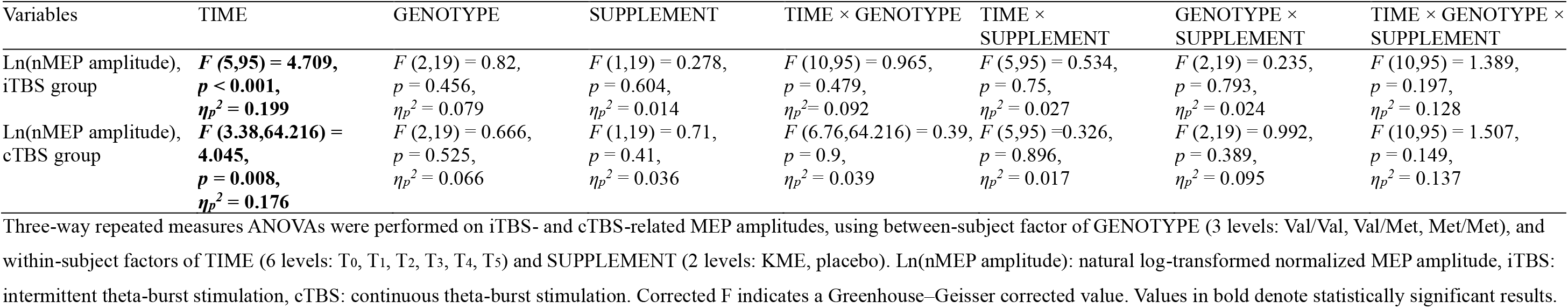
ANOVA results of BDNF genotype interaction with iTBS- and cTBS-related MEP amplitudes.

### 3.5. Effects of KME-TBS on cardiovascular responses

Three-way rmANOVAs of SBP, DBP, MAP, and HR showed a significant main effect of TIME on all values (*p <0.01*). However, the main effect of SUPPLEMENT was significant only in DBP [*F* (1,42) = 7.565, *p* = 0.009, *η_p_^2^* = 0.153] and HR [*F* (1,42) = 145.32, *p* < 0.001, *η_p_^2^* = 0.776]. In addition, there was a significant interaction effect of TIME × SUPPLEMENT [*F* (2.469,103.696) = 24.385, *p* < 0.001, *η_p_^2^* = 0. 367], and TBS × SUPPLEMENT [*F* (1,42) = 8.574, *p* = 0. 005, *η_p_^2^* = 0. 170] on HR. In the DBP analysis, the Bonferroni-adjusted pairwise comparisons for the main effect of SUPPLEMENT showed that DBP significantly decreased after KME in comparison to placebo (mean difference = 2.03; 95% CI, 0.541–3.52; p = 0.009). In the HR analysis, Bonferroni-adjusted pairwise comparisons for the main effect of SUPPLEMENT showed that HR was significantly higher in the KME group in comparison to the placebo group (mean difference = 8.545; 95% CI, 7.115–9.976; p < 0.001). Bonferroni adjusted pairwise comparisons for the interaction effect of TIME × SUPPLEMENT showed that HR in the placebo group significantly increased at T_10min_ in comparison to T_pre-sup_ (mean difference = 2.082; 95% CI, 0.98–4.065; *p* = 0.034); while in the KME group, HR significantly increased at T_post-sup_ in comparison to T_pre-sup_ (mean difference = 6.685; 95% CI, 3.604– 9.765; *p* < 0.001), at T_10min_ in comparison to T_pre-sup_ (mean difference = 6.828; 95% CI, 3.161–10.495; *p* < 0.001), at T20min in comparison to T_pre-sup_ (mean difference = 6.482; 95% CI, 2.685–10.278; *p* < 0.001), and at T30min in comparison to T_pre-sup_ (mean difference = 7.739; 95% CI, 4.015–11.464; *p* < 0.001); furthermore, HR at T_post-sup_ was significantly higher in the KME group compared to the placebo group (mean difference = 10.219; 95% CI, 8.377–12.061; *p* < 0.001), HR at T_10min_ was significantly higher in the KME group compared to the placebo group (mean difference = 10.835; 95% CI, 9.175–12.496; *p* < 0.001), HR at T_20min_ was significantly higher in the KME group compared to the placebo group (mean difference = 10.33; 95% CI, 8.392–12.267; *p* < 0.001), and HR at T_30min_ was significantly higher in the KME group compared to the placebo group (mean difference = 9.417; 95% CI, 7.344–11.489; *p* < 0.001). Bonferroni-adjusted pairwise comparisons for the interaction effect of TBS × SUPPLEMENT showed that the HR increase after KME was significantly greater in the iTBS group than in the cTBS group (mean difference = 10.621; 95% CI, 8.598–12.644; *p* < 0.001 in the iTBS group vs. mean difference = 6.47; 95% CI, 4.447–8.493; *p* < 0.001 in the cTBS group).

Absolute values and full ANOVA results of the cardiovascular measures are shown in Table 8 and Table 9, respectively. The time-course changes in cardiovascular measures are illustrated in Fig. 7.

**Fig. 7.**
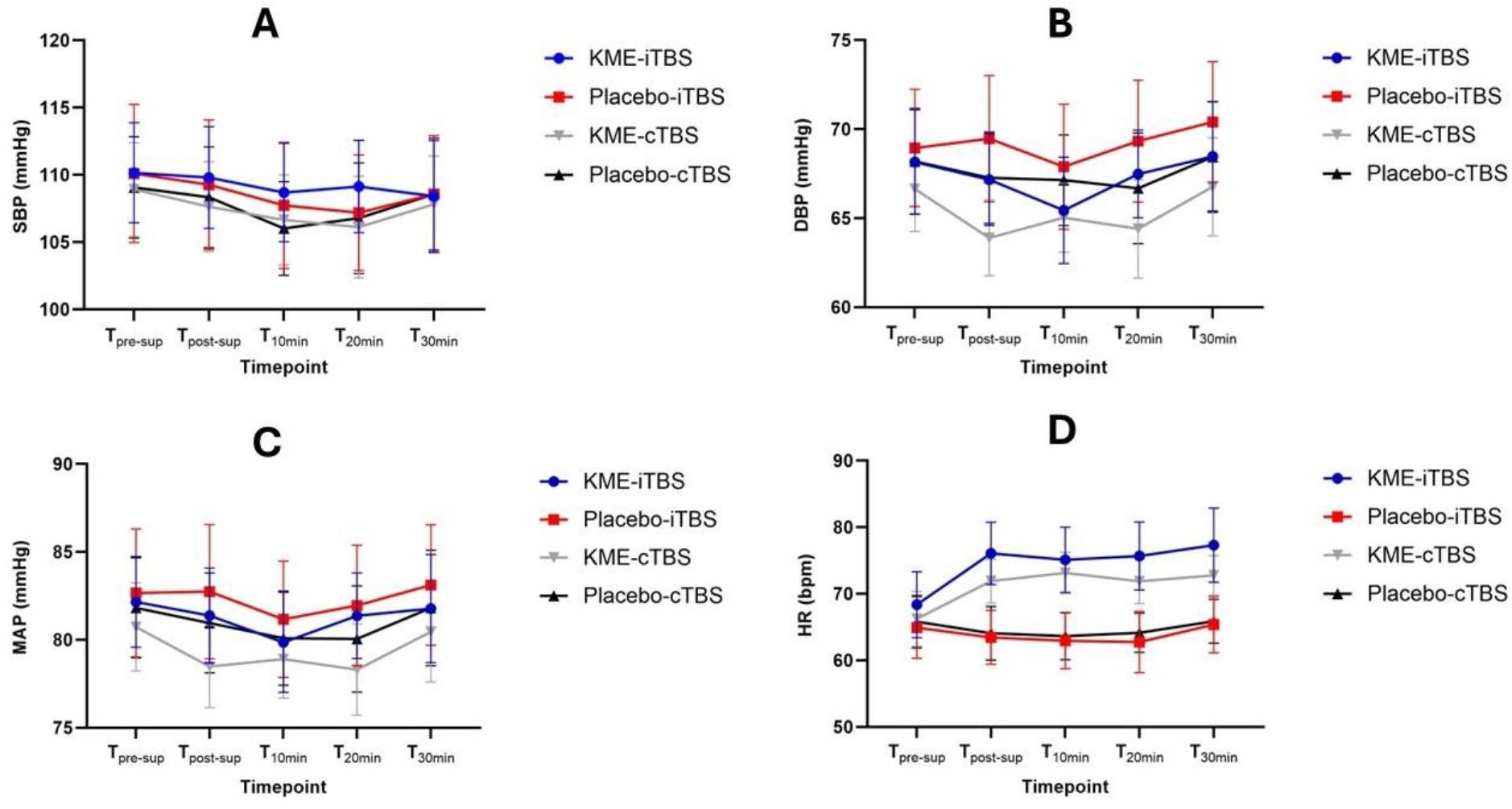
Comparison of cardiovascular responses between the different supplement-TBS combinations. Systolic blood pressure (A), diastolic blood pressure (B), mean arterial pressure (C), and heart rate (D) plotted at every timepoint at baseline (T_pre-sup_), after 1h of supplementation (T_post-sup_), the first 10 minutes post-TBS (T_10min_), the second 10 minutes post-TBS (T_20min_), and the third 10 minutes post-TBS (T_30min_). Error bars denote 95 % confidence interval around the mean.

**Table 8.** Absolute values of cardiovascular measures.

| Variables | iTBS group<br>(N = 22) |  | cTBS group<br>(N = 22) |  |
| --- | --- | --- | --- | --- |
|  | KME | Placebo | KME | Placebo |
| **SBP—mmHg** |  |  |  |  |
| T<sub>pre-sup</sub> | 110.16 ± 8.41 | 110.11 ± 11.59 | 108.92 ± 7.83 | 109.08 ± 8.51 |
| T<sub>post-sup</sub> | 109.81 ± 8.52 | 109.27 ± 10.87 | 107.64 ± 7.56 | 108.34 ± 8.46 |
| T<sub>10min</sub> | 108.69 ± 8.27 | 107.73 ± 10.57 | 106.66 ± 7.54 | 106.02 ± 7.84 |
| T<sub>20min</sub> | 109.14 ± 7.75 | 107.2 ± 9.7 | 106.13 ± 8.53 | 106.79 ± 9.26 |
| T<sub>30min</sub> | 108.42 ± 9.34 | 108.57 ± 9.8 | 107.83 ± 8.05 | 108.58 ± 9.37 |
| **DBP—mmHg** |  |  |  |  |
| T<sub>pre-sup</sub> | 68.16 ± 6.6 | 68.95 ± 7.43 | 66.65 ± 5.39 | 68.2 ± 6.7 |
| T<sub>post-sup</sub> | 67.17 ± 5.82 | 69.47 ± 8.01 | 63.9 ± 4.81 | 67.27 ± 5.79 |
| T<sub>10min</sub> | 65.45 ± 6.73 | 67.89 ± 7.91 | 65.03 ± 4.37 | 67.14 ± 5.73 |
| T<sub>20min</sub> | 67.49 ± 5.57 | 69.33 ± 7.73 | 64.4 ± 6.22 | 66.68 ± 7.03 |
| T<sub>30min</sub> | 68.46 ± 6.91 | 70.4 ± 7.64 | 66.77 ± 6.21 | 68.44 ± 7.02 |
| **MAP—mmHg** |  |  |  |  |
| T<sub>pre-sup</sub> | 82.16 ± 5.81 | 82.67 ± 8.2 | 80.74 ± 5.65 | 81.82 ± 6.41 |
| T<sub>post-sup</sub> | 81.39 ± 6.09 | 82.74 ± 8.62 | 78.48 ± 5.27 | 80.96 ± 6.42 |
| T<sub>10min</sub> | 79.86 ± 6.41 | 81.17 ± 7.46 | 78.91 ± 4.99 | 80.1 ± 6.04 |
| T<sub>20min</sub> | 81.37 ± 5.48 | 81.95 ± 7.73 | 78.31 ± 5.86 | 80.05 ± 6.81 |
| T<sub>30min</sub> | 81.78 ± 6.93 | 83.12 ± 7.73 | 80.46 ± 6.43 | 81.82 ± 7.42 |
| **HR—bpm** |  |  |  |  |
| T<sub>pre-sup</sub> | 68.37 ± 11.17 | 64.93 ± 10.43 | 66.24 ± 9.3 | 65.83 ± 8.88 |
| T<sub>post-sup</sub> | 76.07 ± 10.55 | 63.45 ± 9.14 | 71.91 ± 7.41 | 64.09 ± 9.07 |
| T<sub>10min</sub> | 75.11 ± 11.05 | 62.94 ± 9.43 | 73.16 ± 6.92 | 63.65 ± 7.96 |
| T<sub>20min</sub> | 75.68 ± 11.52 | 62.75 ± 10.39 | 71.89 ± 7.5 | 64.16 ± 6.62 |
| T<sub>30min</sub> | 77.32 ± 12.54 | 65.37 ± 9.5 | 72.77 ± 6.66 | 65.88 ± 7.43 |

Values are means ± standard deviation. SBP: systolic blood pressure, DBP: diastolic blood pressure, MAP: mean arterial pressure, HR: heart rate, iTBS: intermittent theta-burst stimulation, cTBS: continuous theta-burst stimulation, KME: ketone monoester.

**Table 9.**
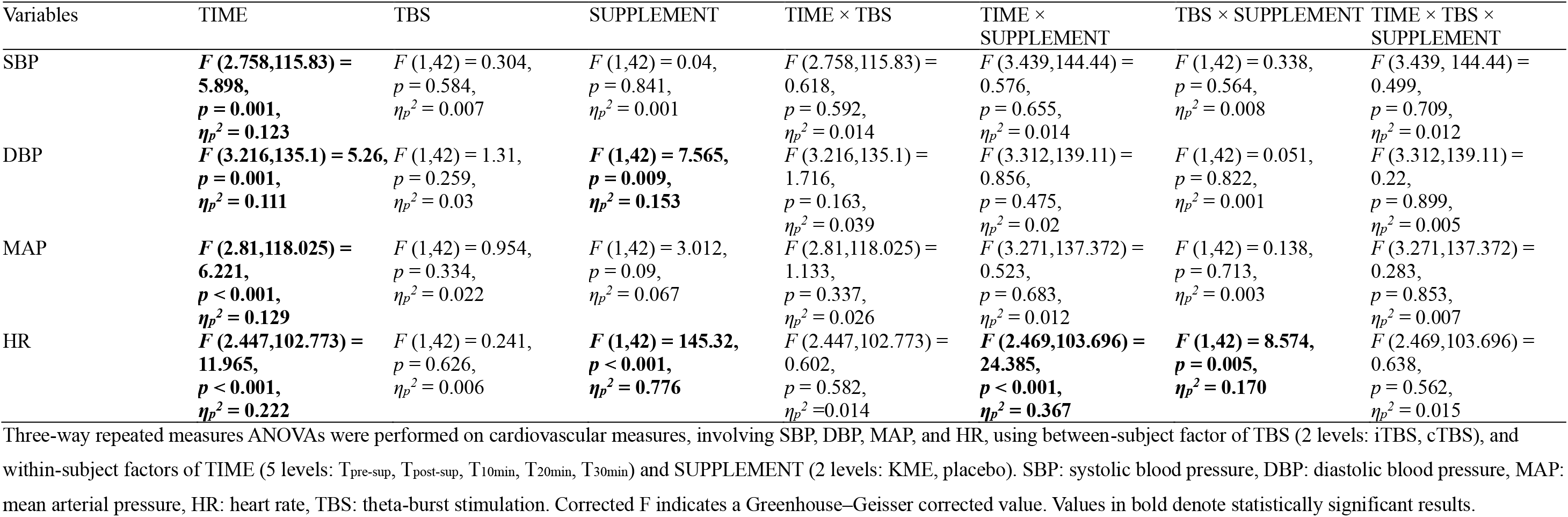
ANOVA results of cardiovascular measures.

### 3.6. Effects of KME-TBS on blood biomarkers

Three-way rmANOVAs of blood BHB showed a significant effect of TIME [*F* (1,42) = 559.981, *p* < 0.001, *η_p_^2^* = 0.93], SUPPLEMENT [*F* (1,42) = 581.871, *p* < 0.001, *η_p_^2^* = 0.933], and TIME × SUPPLEMENT [*F* (1,42) = 594.796, *p* < 0.001, *η_p_^2^* = 0.934]. Bonferroni adjusted pairwise comparisons for the interaction effect of TIME × SUPPLEMENT showed that blood BHB in the placebo group significantly increased at B_2_ in comparison to B_1_ (mean difference = 0.055; 95% CI, 0.007–0.102; *p* = 0.024); while in the KME group, BHB significantly increased at B_2_ in comparison to B_1_ (mean difference = 3.398; 95% CI, 3.116–3.679; *p* < 0.001); furthermore, blood BHB at B_2_ was significantly higher in the KME group compared to the placebo group (mean difference = 3.364; 95% CI, 3.092–3.635; *p* < 0.001), indicating a successful induction of ketosis as planned (Fig. 8).

**Fig. 8.**
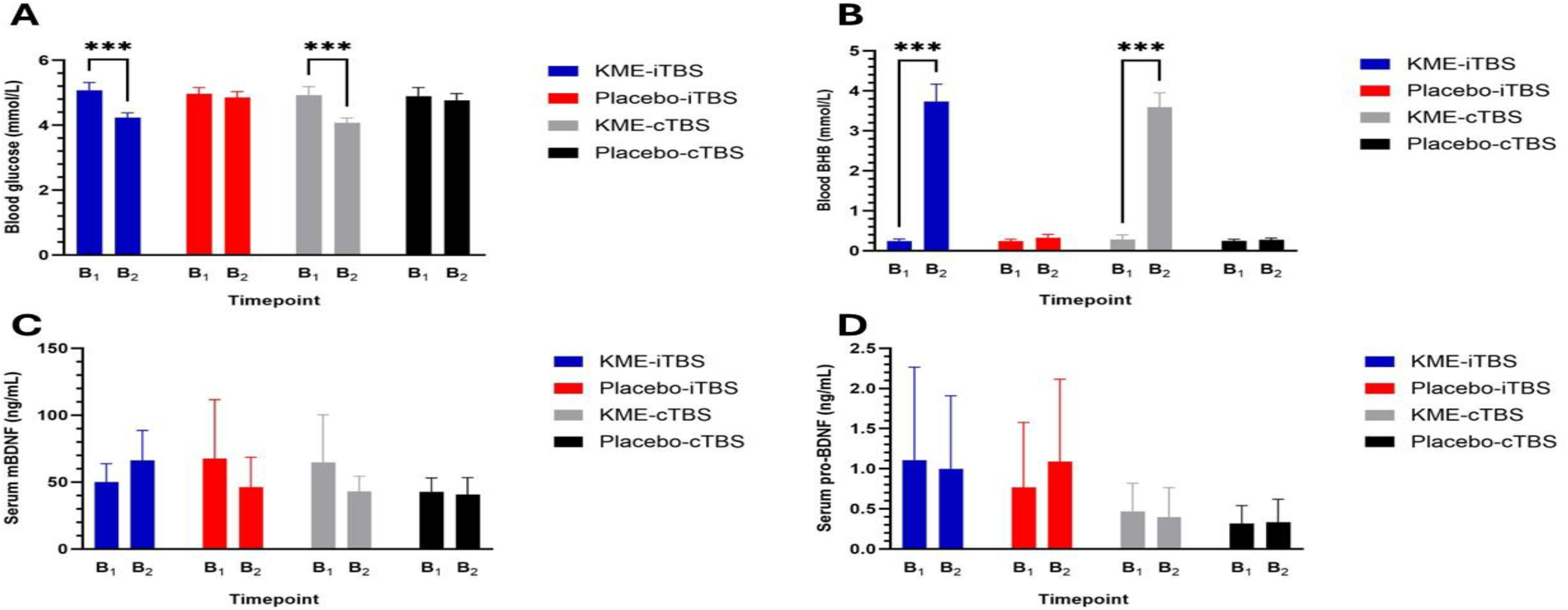
Comparison of blood biomarkers between the different supplement-TBS combinations. Blood glucose (A), Blood BHB (B), Serum mBDNF (C), and Serum pro-BDNF (D) at baseline (B_1_) and the end of the session (B_2_). Error bars denote 95% confidence interval around the mean. (***) denotes *p* < 0.001.

Three-way rmANOVAs of blood glucose showed a significant effect of TIME [*F* (1,42) = 59.521, *p* < 0.001, *η_p_^2^* = 0.586], SUPPLEMENT [*F* (1,42) = 17.174, *p* < 0.001, *η_p_^2^* = 0.29], and TIME × SUPPLEMENT [*F* (1,42) = 47.61, *p* < 0.001, *η_p_^2^*= 0.531]. Bonferroni-adjusted pairwise comparisons for the interaction effect of TIME × SUPPLEMENT showed that blood glucose in the KME group significantly decreased at B_2_ in comparison to B_1_ (mean difference = 0.848; 95% CI, 0.665–1.030; *p* < 0.001); furthermore, blood glucose at B_2_ was significantly lower in the KME group compared to the placebo group (mean difference = 0.655; 95% CI, 0.509–0.800; *p* < 0.001).

For serum mBDNF and pro-BDNF, non-parametric Wilcoxon signed-rank tests revealed no statistically significant pre-to-post changes within any group (Table 11).

**Table 10.** Absolute values of Blood biomarkers.

| Variables | iTBS group<br>(N = 22) |  | cTBS group<br>(N = 22) |  |
| --- | --- | --- | --- | --- |
|  | KME | Placebo | KME | Placebo |
| Blood glucose—<br>mmol/L |  |  |  |  |
| B<sub>1</sub> | 5.07 ± 0.55 | 4.96 ± 0.45 | 4.93 ± 0.59 | 4.9 ± 0.61 |
| B<sub>2</sub> | 4.23 ± 0.34 | 4.85 ± 0.42 | 4.07 ± 0.34 | 4.76 ± 0.48 |
| Blood BHB—mmol/L |  |  |  |  |
| B<sub>1</sub> | 0.25 ± 0.11 | 0.24 ± 0.11 | 0.29 ± 0.25 | 0.25 ± 0.09 |
| B<sub>2</sub> | 3.73 ± 0.99 | 0.33 ± 0.19 | 3.6 ± 0.8 | 0.27 ± 0.1 |
| Serum mBDNF—<br>ng/mL |  |  |  |  |
| B<sub>1</sub> | 38.84 (32.63 – 50.72) | 32.59 (28.27 – 55.46) | 38.55 (27.88 – 51.78) | 38.95 (31.77 – 47.79) |
| B<sub>2</sub> | 48.89 (30.06 – 96.17) | 34.75 (28.32 – 38.96) | 36.33 (31.23 – 43.12) | 33.75 (24.29 – 62.11) |
| Serum pro-BDNF—<br>ng/mL |  |  |  |  |
| B<sub>1</sub> | 0.35 (0.09 – 2.57) | 0.38 (0.17 – 1.78) | 0.28 (0.16 – 0.95) | 0.19 (0.08 – 0.58) |
| B<sub>2</sub> | 0.46 (0.09 – 2.52) | 0.38 (0.09 – 1.65) | 0.33 (0.16 – 0.94) | 0.23 (0.09 – 0.6) |

Values are means ± standard deviation for normally distributed data and medians (interquartile range) for non-normally distributed data.

**Table 11.** Pairwise comparison of serum BDNF levels.

| Comparisons | iTBS group<br>(N = 22) |  | cTBS group<br>(N = 22) |  |
| --- | --- | --- | --- | --- |
|  | KME | Placebo | KME | Placebo |
| Serum mBDNF |  |  |  |  |
| B<sub>2</sub> vs. B<sub>1</sub> | $T = 59, p = 0.117$ | $T = 24, p = 0.424$ | $T = 28, p = 0.069$ | $T = 52, p = 0.408$ |
| Serum pro-BDNF |  |  |  |  |
| B<sub>2</sub> vs. B<sub>1</sub> | $T = 22, p = 0.953$ | $T = 6, p = 0.345$ | $T = 10, p = 0.5$ | $T = 21, p = 0.237$ |

Serum BDNF levels were analyzed using the non-parametric Wilcoxon matched-pair signed-rank test.

**Table 12.** ANOVA results of blood glucose and BHB.

| Variables | TIME | TBS | SUPPLEMENT | TIME × TBS | TIME × SUPPLEMENT | TBS × SUPPLEMENT | TIME × TBS × SUPPLEMENT |
| --- | --- | --- | --- | --- | --- | --- | --- |
| Glucose | $F(1,42) = 59.521$,<br>$p < 0.001$,<br>$\eta_p^2 = 0.586$ | $F(1,42) = 1.387$,<br>$p = 0.246$,<br>$\eta_p^2 = 0.032$ | $F(1,42) = 17.174$,<br>$p < 0.001$,<br>$\eta_p^2 = 0.290$ | $F(1,42) = 0.016$,<br>$p = 0.9$,<br>$\eta_p^2 = 0.001$ | $F(1,42) = 47.61$,<br>$p < 0.001$,<br>$\eta_p^2 = 0.531$ | $F(1,42) = 0.283$,<br>$p = 0.597$,<br>$\eta_p^2 = 0.007$ | $F(1,42) = 0.001$,<br>$p = 0.983$,<br>$\eta_p^2 = 0.001$ |
| BHB | $F(1,42) = 559.981$,<br>$p < 0.001$,<br>$\eta_p^2 = 0.930$ | $F(1,42) = 0.243$,<br>$p = 0.624$,<br>$\eta_p^2 = 0.006$ | $F(1,42) = 581.871$,<br>$p < 0.001$,<br>$\eta_p^2 = 0.933$ | $F(1,42) = 0.682$,<br>$p = 0.414$,<br>$\eta_p^2 = 0.016$ | $F(1,42) = 594.796$,<br>$p < 0.001$,<br>$\eta_p^2 = 0.934$ | $F(1,42) = 0.032$,<br>$p = 0.859$,<br>$\eta_p^2 = 0.001$ | $F(1,42) = 0.172$,<br>$p = 0.681$,<br>$\eta_p^2 = 0.004$ |

Three-way repeated measures ANOVAs were performed on blood glucose and BHB using between-subject factor of TBS (2 levels: iTBS, cTBS), and within-subject factors of TIME (2 levels: B<sub>1</sub>, B<sub>2</sub>) and SUPPLEMENT (2 levels: KME, placebo). BHB: β-hydroxybutyrate, TBS: theta-burst stimulation. Values in bold denote statistically significant results.

### 3.7. Association between corticospinal excitability, cardiovascular response, and blood biomarkers

Exploratory correlation analyses revealed that in the KME group (N = 44), blood levels of BHB at B_2_ significantly correlated with mBDNF (rho = 0.355, *p* =0.046). In addition, BHB at B_2_ significantly correlated with heart rate across all timepoints, at T_post-sup_ (r = 0.366, p = 0.015), T_10min_ (*r* = 0.336, *p* = 0.026), T_20min_ (*r* = 0.430, *p* = 0.004), and T_30min_ (r = 0.359, p = 0.017), indicating that the heart rate acceleration was dose-dependent on KME (Fig. 9).

**Fig. 9.**
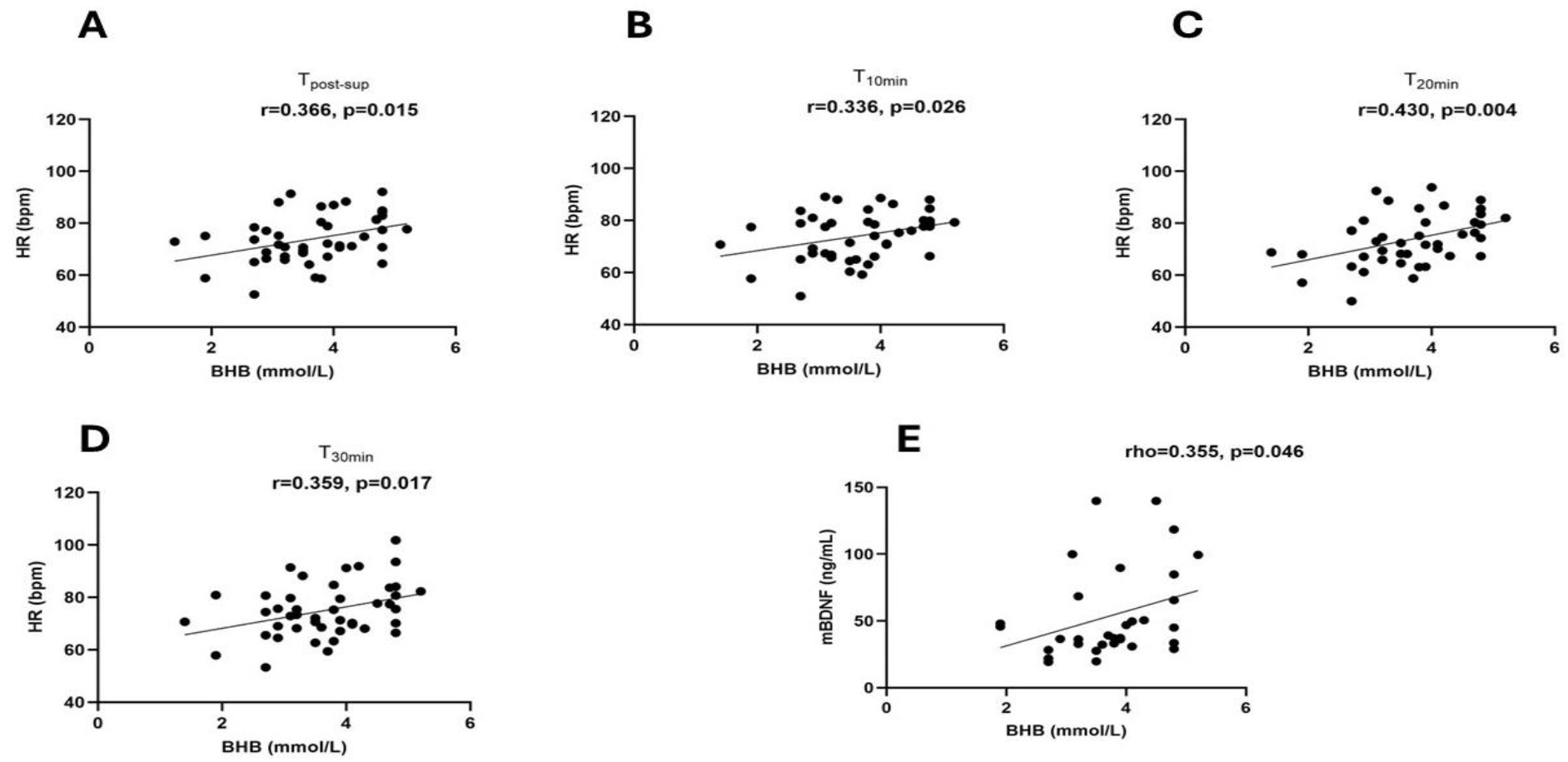
Significant associations with blood BHB levels, total sample (N=44). Grouped scatterplots illustrating the correlation between blood BHB levels with heart rate at T_post-sup_ (A), T_10min_ (B), T_20min_ (C), T_30min_ (D), and serum mBDNF (E). The single regression line represents the pooled cohort trend.

In the KME-cTBS group (N = 22), there was a significant correlation between blood BHB at B_2_ and normalized MEP amplitude at T_2_ (rho = 0.554, *p* = 0.008), T_3_ (rho = 0.512, *p* = 0.015), and nearly significant correlation with the MEP grand average (rho = 0.418, *p* = 0.053) as shown in Fig. 10.

**Fig. 10.**
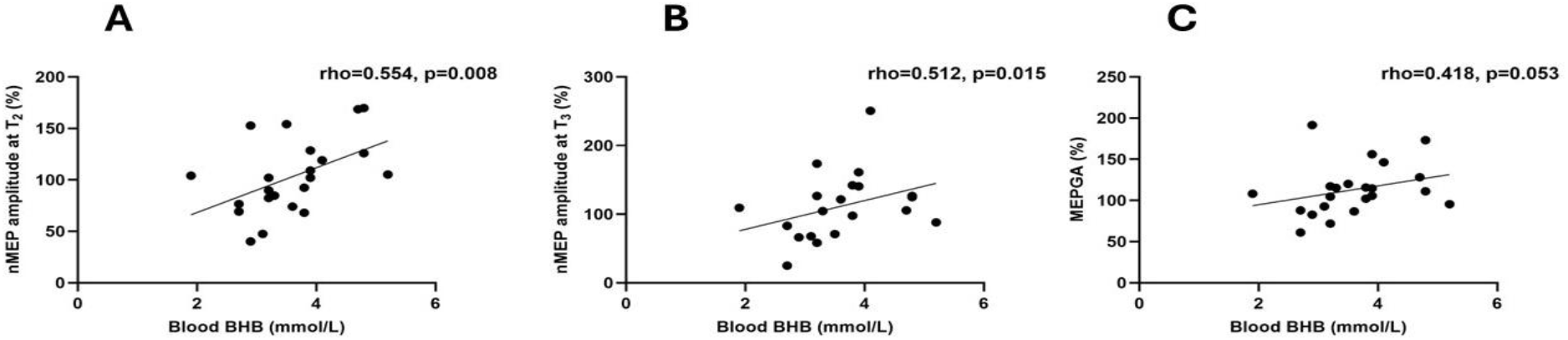
Association between blood BHB and MEP modulation in the KME-cTBS group. Bivariate scatterplots illustrating the association between blood BHB and normalized MEP amplitudes, i.e., % change to baseline, at T_2_ (A), T_3_ (B), and MEPGA (C).

In the placebo-iTBS group (N = 22), there was a significant correlation between blood glucose at B_2_ and normalized MEP amplitude at T_2_ (rho = −0.583, *p* = 0.004), T_3_ (rho = −0.447, *p* = 0.037), T_5_ (rho = −0.537, *p* = 0.01), and MEP grand average (rho = −0.530, *p* = 0.011) as shown in Fig.11. Collectively, this might indicate a differential utilization of blood metabolites by the brain depending on the TBS protocol.

**Fig. 11.**
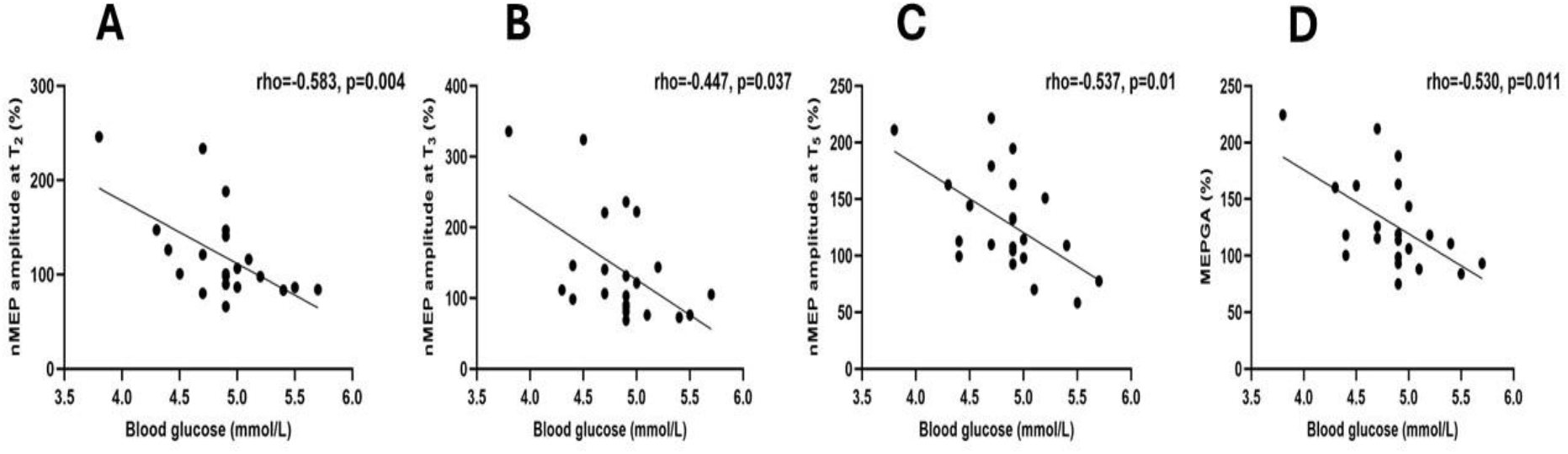
Association between blood glucose and MEP modulation in the placebo-iTBS group. Bivariate scatterplots illustrating the association between blood glucose and normalized MEP amplitudes, i.e., % change to baseline, at T_2_ (A), T_3_ (B), T_5_ (C), and MEPGA (D).

## 4. Discussion

This study provides the first evidence that the combination of KME, a fast method to induce nutritional ketosis, with TBS, a fast TMS paradigm to induce neuroplasticity, is feasible, well-tolerated, and has concurrent neurological and cardiovascular effects. This adds to the growing body of literature that highlights the multisystem impact of ingested ketones on the human body.

### 4.1. Effects of KME on TBS-induced plasticity and baseline corticospinal excitability

The primary hypothesis was not supported. KME did not significantly modify the overall TBS-induced MEPs. Exploratory within-condition analyses showed a blunted MEP modulation in the KME-cTBS condition only, although the supplement-related interaction was not significant. This pattern raises the possibility that KME influences cTBS responses, but the absence of a significant supplement interaction precludes a definitive conclusion. Noteworthy, the original hypothesis was merely translational in nature. In other words, we aimed to translate pre-clinical studies, which have unanimously shown that the ketone body BHB upregulates the neuronal BDNF gene expression via multiple molecular pathways (Giacco et al., 2026; Hu et al., 2020; Hu et al., 2018; Kwak et al., 2021; Marosi et al., 2016; Sleiman et al., 2016; Sun et al., 2022; Trotta et al., 2022; Zhang et al., 2022). BDNF, in turn, has an indispensable role in cellular neuroplasticity induction, with mBDNF inducing long-term potentiation (LTP) plasticity and pro-BDNF inducing long-term depression (LTD) plasticity, a principle known as the ‘yin and yang’ BDNF effect (Lu et al., 2005; Pisani et al., 2023; Woo et al., 2005). This duality of mBDNF/pro-BDNF function in synaptic plasticity is believed to drive the duality of iTBS/cTBS after-effects (Brunoni et al., 2008; C. W. Lee et al., 2023). Admittedly, however, the human brain is vastly more complex than isolated preclinical models. This explains why preclinical breakthroughs do not always translate into the clinical field (Seyhan, 2019), a bottleneck that is particularly pronounced in brain-related research.

Within the iTBS cohort, we found that KME did not have a significant effect on iTBS-induced plasticity, at least during the 30 minutes post-iTBS. Hence, the iTBS protocol successfully induced M1 plasticity, significantly increasing MEP amplitudes by a similar magnitude regardless of KME or placebo supplementation.

Within the cTBS cohort, however, significant increases in MEP amplitudes were observed at T_4_ and T_5_ during the placebo session, whereas MEP amplitudes did not significantly differ from baseline at any post-cTBS time point during the KME session. This exploratory pattern may indicate attenuation of the observed post-cTBS modulation after KME priming. Nevertheless, the lack of significant supplement-related interaction means that a differential KME effect cannot be concluded from these condition-specific comparisons alone. We discuss this observation from two distinct perspectives.

First, the cTBS protocol unexpectedly increased MEP amplitudes in the placebo-cTBS condition. This stands in sharp contrast to the seminal work by Huang et al., who reported a significant suppressing effect of the cTBS protocol on MEP amplitudes (Huang et al., 2005). The authors of that study recruited only 9 participants, applied the cTBS protocol at an intensity of 80% active motor threshold (AMT), and used a flat coil connected to a Magstim stimulator. In contrast, we recruited 22 subjects in our cTBS protocol, providing a relatively higher statistical power (Button et al., 2013), applied the cTBS protocol at 70% RMT intensity, reducing the confounding effect of muscle contraction during AMT (Gentner et al., 2008; Goldsworthy et al., 2014; Iezzi et al., 2008), and used a 150°-bent Cool-B70 coil connected to a MagVenture stimulator. Crucially, the MagVenture biphasic waveform features an opposite current direction compared to the Magstim stimulator (Corp et al., 2021; Kanig et al., 2025). Additionally, the 150°-bent Cool-B70 coil has a greater penetration depth and stronger magnetic field gradient than the standard flat coils (Deng et al., 2013; Drakaki et al., 2022; MagVenture, 2024). Arguably, therefore, our distinct combination of hardware configuration and current orientation might have recruited a different population of cortical neurons than those targeted in the original cTBS protocol configuration (Huang et al., 2005; Suppa et al., 2016). Moreover, there have been calls recently challenging the excitatory-inhibitory dichotomy dogma of the iTBS and cTBS protocols (Hermiller, 2026; Hussain & Freedberg, 2025; Yassi et al., 2026). These perspectives emphasize that no TBS protocol is inherently excitatory or inhibitory by nature, but rather determined by the neural context, wherein specific technical hardware configurations interact with distinct target circuit properties and ongoing functional states (Hermiller, 2026; Hussain & Freedberg, 2025; Yassi et al., 2026).

The second angle of our perspective pertains to the suppressing effect of the KME supplement on cTBS-induced plasticity. Notably, circulating mBDNF and pro-BDNF concentrations did not change significantly, and the primary BDNF-mediated hypothesis was therefore not supported by the peripheral biomarker data. Still, we cannot exclude local neurotrophic signaling within the M1, particularly given the limited sampling time points and missing biomarker measurements. On the other hand, all participants were genotyped for the rs6265 variant of the BDNF gene, allowing us to control for its potential confounding effect during analysis. With neurotrophic changes being less likely to explain the reduced cTBS effects after KME supplementation, the observed results may reflect an altered neurotransmission.

Pharmacological studies have shown that the drug memantine, an NMDA receptor antagonist, blocked cTBS after-effects, indicating the dependence of cTBS-induced plasticity on glutamatergic neurotransmission (Huang et al., 2007). On the other hand, an earlier neuroimaging MRS study found that cTBS increased regional GABA concentrations when stimulating the M1 cortex (Stagg et al., 2009). A subsequent MRS study found that cTBS simultaneously increased GABA concentrations in the stimulated left hemisphere and decreased GABA in the unstimulated right hemisphere (Matsuta et al., 2022). However, cTBS did not have any effects on the GABA levels when stimulating the visual cortex (Stoby et al., 2022), suggesting regional differences in the cTBS effect on GABA. On the other hand, while extended ketogenic diets are a well-known antiepileptic intervention driven mainly by higher GABA/glutamate ratio (Pflanz et al., 2019; Qiao et al., 2024), such effects do not seem to extend to the acute nutritional ketosis induced by KME. In fact, MRS imaging showed a significant increase in glutamate (but not GABA) in the occipital cortex after acute KME supplementation (Steinwurzel et al., 2025). Other MRS studies revealed that acute KME supplementation significantly decreased both glutamate and GABA concentrations in the cingulate cortex (Hone-Blanchet et al., 2023; van Nieuwenhuizen et al., 2025). Taken together, the GABA/glutamate system seems to be a plausible point of convergence between the cTBS protocol and KME. However, whether KME reduced the cTBS-induced plasticity by modulating GABA or glutamate in the left motor cortex remains unknown and subject to further investigation. Additionally, alternative mechanisms underlying KME-TBS interaction cannot be ruled out.

Finally, KME did not modulate the baseline corticospinal excitability indices, namely RMT and MEP amplitude and latency, indicating that acute ketone elevation did not alter baseline excitability when the motor cortex was at rest. Rather, the KME exerted neuromodulatory effects that required state-dependent synaptic challenge, such as cTBS.

### 4.2. Effects of KME on cardiovascular responses

The KME supplement significantly increased heart rate and decreased diastolic blood pressure in comparison to placebo. Several studies reported an increase in resting heart rate post-KME supplementation (Oneglia et al., 2023; Selvaraj et al., 2022; Selvaraj et al., 2025). This is mediated by a combination of decreased cardiac vagal modulation (Thiessen et al., 2026) along with a hyperdynamic cardiac state characterized by an increased left ventricular ejection fraction (Berg-Hansen et al., 2024; Selvaraj et al., 2022). The latter effect in particular is driving a rising interest in the cardiology field. Recent randomized clinical trials have demonstrated beneficial effects of KME among patients with heart failure with reduced ejection fraction (Berg-Hansen et al., 2024), type 2 diabetes and heart failure with preserved ejection fraction (Gopalasingam et al., 2024), and cardiogenic shock (Berg-Hansen et al., 2023). The primary point of alignment across these trials is that exogenous ketones through KME act as potent metabolic inotropes that significantly boost cardiac performance. The lack of change in circulating metanephrines after KME ingestion indicates that exogenous ketones act as metabolic inotropes, augmenting heart contractility through energetics rather than traditional sympathetic stimulation (Berg-Hansen et al., 2025). This is crucial as the enhancement in cardiac output occurs without a parallel increase in myocardial oxygen consumption in the diseased heart (Elliott et al., 2026).

On the other hand, the effect of KME on diastolic blood pressure is less pronounced in the literature, with most studies reporting non-significant changes (Marcotte-Chenard et al., 2026; Marcotte-Chenard et al., 2023; McClure et al., 2025; Seto et al., 2025). However, the present study may offer greater statistical sensitivity to detect subtle changes, owing to the larger sample size (N = 44 compared to N < 30) and the crossover design. Crossover designs are well known to reduce the inter-individual variability in responses, as every individual serves as his or her own control. This in turn allows for the detection of smaller effect sizes and reduces the variation in nonspecific (nontreatment-related) factors (Stoney & Johnson, 2018).

### 4.3. Influence of BDNF genetic polymorphism on the TBS-induced neuroplasticity

The BDNF rs6265 single nucleotide polymorphism had no interaction with MEP amplitude modulation at any timepoint across any supplement-TBS combination. The seminal study that reported a differential response to iTBS and cTBS depending on the BDNF genotype was published by Cheeran and colleagues in 2008 (Cheeran et al., 2008). Across these two decades, subsequent studies have reported conflicting results regarding the interaction between BDNF genotypes and TBS-induced plasticity in the motor cortex. A recent systematic review pooling data from 36 TMS studies overall reported that only 53% of studies in healthy individuals demonstrated significant BDNF genotype-dependent differences in neuroplasticity (Kuo et al., 2026). Notably, the studies that reported non-significant MEP findings had larger sample sizes and categorized their participants into three groups (Val/Val, Val/Met, and Met/Met), similar to our approach. In contrast, studies that reported significant findings had smaller sample and merged participants with Val/Met and Met/Met genotypes as Met carriers to analyze their data as two groups (Val/Val vs. Met carriers), due to the small sample size and the assumption that Val/Met and Met/Met exhibit similar phenotypic profiles (Egan et al., 2003). This merging approach, however, may have affected the judgment of the genotype-related changes in MEP amplitudes as suggested by a former systematic review (Sasaki et al., 2021).

### 4.4. Effect of KME-TBS on serum BDNF levels

We found that serum levels of pro-BDNF and mBDNF did not significantly change in any supplement-TBS combination. However, we acknowledge that the optical density values fell outside the standard curve in 55% of pro-BDNF samples (below the curve) and 26% of mBDNF samples (above the curve) during ELISA and were therefore excluded from statistical analyses. The first study that reported the measurability of pro-BDNF in human serum was the one by Yoshida et al. (Yoshida et al., 2012). The authors acknowledged the low serum concentrations of pro-BDNF among healthy adults, with 15/40 subjects having serum levels below the minimum detectable range of the kit (Yoshida et al., 2012). Notably, the majority of commercial immunoassays used to analyze peripheral BDNF concentrations in the literature do not differentiate between the precursor protein (pro-BDNF) and the mature form (mBDNF), despite their having opposite functional roles in neuroplasticity. This is a major cause of heterogeneity when interpreting “peripheral BDNF” results. Yet, only a handful of studies have raised this issue (Balietti et al., 2018; Lim et al., 2015; Polacchini et al., 2015). To add to the level of complexity, mBDNF levels in the serum and plasma did not show any correlation (Gejl et al., 2019), indicating that they reflect two different pools of the protein. Therefore, the high cross-reactivity of commercial kits between pro-BDNF and mBDNF, along with the different mBDNF levels in serum and plasma, needs to be taken into account when interpreting "peripheral BDNF levels" across the literature. Nonetheless, a study on ketogenic oils in healthy older adults reported a distinct response pattern based on the level of ketosis. Serum pro-BDNF increased more significantly during a high level of ketosis compared to a low level and was predictable by the blood BHB values (Norgren et al., 2021). On the other hand, we found a significant positive correlation between the blood ketone BHB and serum mBDNF levels. However, unlike the study by Norgren et., we administered a fixed dose of the ketogenic supplement (500 mg/kg) and recruited younger subjects. It is well-established that the neurological effects of ketones differ significantly based on age (Antal et al., 2025; Hone-Blanchet et al., 2023). A study on KME reported a significant increase in plasma BDNF after an oral glucose tolerance test in obese subjects (Walsh et al., 2020), arguing for the neuroprotective potential of KME. A subsequent study by the same group on patients with type 2 diabetes demonstrated that KME did not alter serum or plasma BDNF levels during fasting (Baranowski et al., 2025), arguing that KME may not directly upregulate BDNF expression under basal conditions but rather provide a neuroprotective buffer during hyperglycemic spikes. On the other hand, TBS studies have often investigated circulating BDNF changes after multiple TBS sessions for treatment, with conflicting results (Ankit et al., 2022; Rashid-López et al., 2026; Sanna et al., 2020). Nonetheless, there is limited evidence regarding the effect of a single TBS session on serum mBDNF and pro-BDNF isoforms.

### 4.5. Effect of KME-TBS on blood glucose and BHB

As expected, blood levels of the ketone body BHB significantly increased after KME compared to placebo. This indicates that the target ketosis was successfully achieved as planned. The mean level of blood BHB was > 3 mmol/L, which is sufficient to deliver BHB through the blood-brain barrier (Hasselbalch et al., 1996; Mikkelsen et al., 2015; van Nieuwenhuizen et al., 2025).

In addition, we found a significant decrease in blood glucose after KME compared to placebo. This effect is well-established in the KME literature and is believed to result from an improved insulin sensitivity along with a reduction in hepatic gluconeogenesis (Falkenhain et al., 2022). Moreover, because the system shifts its fuel preference toward burning ketones, peripheral and cerebral glucose utilization naturally drops, reducing the immediate demand for circulating glucose (Hasselbalch et al., 1996; Hone-Blanchet et al., 2023; van Nieuwenhuizen et al., 2025). Therefore, the glucose-lowering effect of KME is garnering attention recently as a potential treatment for diabetes mellitus.

Randomized controlled trials have shown that pre-meal KME supplementation reduced postprandial glucose and lipid concentrations in patients with type 2 diabetes (Bangshaab et al., 2026; Monteyne et al., 2024). In addition, a study found that KME significantly increased cardiac output and reduced systemic vascular resistance but had no effect on glucose, potentially due to the low KME dose (115 mg/kg), in patients with type 2 diabetes (Perissiou et al., 2025). Importantly, extended KME-induced ketosis (by consuming 3-4 KME doses daily for 2-4 weeks) was safe in patients with type 2 diabetes (Falkenhain et al., 2024; Gopalasingam et al., 2024; Soto-Mota et al., 2021), improved their glycemic control (Soto-Mota et al., 2021), and enhanced cardiac function in diabetes-related heart failure (Gopalasingam et al., 2024). This indication seems counterintuitive given that ketosis is generally classified as “evil” in diabetes due to the risk of diabetic ketoacidosis (Kolb et al., 2021). However, the KME ingestion generates a mild state of metabolic ketosis where the BHB level is < 5 mM/L. This is unlike severe metabolic ketosis, e.g., diabetic ketoacidosis, where BHB levels accumulate to more than 20 mM/L due to pathological insulin resistance, followed by a life-threatening decline in blood pH (Kolb et al., 2021; Poff et al., 2020). In addition, the consumption of exogenous ketone bodies through KME is thought to yield negative feedback on the generation of internal ketone bodies. Studies have shown that exogenous ketones bind to the nicotinic acid receptor PUMA-G on adipocytes. This activation directly inhibits lipolysis, which reduces the release of non-esterified fatty acids from adipose tissue (Falkenhain et al., 2022; Taggart et al., 2005). Since these fatty acids are the primary substrate for hepatic ketogenesis, lowering their concentration significantly decreases the production of endogenous ketones, which might buffer against the development of ketoacidosis (Falkenhain et al., 2022; Kolb et al., 2021).

On the other hand, the TBS did not have any effects on blood glucose or BHB on the group level. Remarkably, however, there was a significant positive correlation between blood BHB levels and normalized MEP amplitudes post-cTBS in the KME-cTBS group. Additionally, there was a significant negative correlation between blood glucose levels and normalized MEP amplitudes post-iTBS in the placebo-iTBS group. The mechanistic rationale for this phenomenon is unclear. However, it might indicate a differential utilization of blood metabolites by the brain depending on the TBS protocol (Aceves-Serrano et al., 2022; Kinney & Hanlon, 2022).

### 4.6. Non-specific physiological changes

Finally, we discuss a few physiological changes that occurred in the TIME factor only. First, systolic blood pressure and mean arterial pressure dropped in all sessions over time, without any interaction effect. This indicates that their drop was independent of any intervention, but rather secondary to another factor, most likely the alleviation of anxiety of a new procedure since all subjects were naïve to TMS (Russell & Lightman, 2019). Second, MEP latency prolonged significantly toward the end of all sessions, without any interaction effect. We attribute this MEP latency prolongation to the sleepiness reported by most subjects. This is consistent with the findings of Avesani and colleagues who reported a significant prolongation of MEP latency during sleepiness, probably due to thalamocortical hyperpolarization during sleep onset, which modulates cortical reactivity to sensory inputs (Avesani et al., 2008).

### 4.7. Limitations and Future Recommendations

A number of factors limit the generalizability of our findings and should be acknowledged. First, our neurophysiological outcome measures depended solely on the single-pulse elicited MEPs, which may not have captured the full neurophysiological effects of KME on the brain. Future studies should incorporate additional outcome measures, such as TMS-EEG and paired-pulse TMS protocols. Second, our outcome measures were confined to the first 30 minutes post-TBS. Therefore, the duration of the aftereffects of KME-TBS combinations remains an open question. Third, we established the feasibility of KME and iTBS/cTBS protocols in healthy young adults. However, the clinical relevance of our findings in patient populations remains to be explored.

## 5. Conclusions

In the present study, we demonstrated for the first time that the combination of KME, a fast method to induce nutritional ketosis, with TBS, a fast TMS paradigm to induce neuroplasticity, is feasible, well-tolerated, and has concurrent neurological and cardiovascular effects. This adds to the growing body of literature that highlights the multisystem impact of ingested ketones on the human body. In particular, acute KME supplementation reliably induced ketosis, reduced blood glucose, increased heart rate and was associated with lower diastolic blood pressure in healthy young adults. KME did not alter resting corticospinal excitability and did not significantly modify the overall TBS-induced MEPs in the primary factorial analysis. Exploratory condition-specific analyses showed late MEP facilitation after placebo-cTBS but no significant post-cTBS modulation during the KME condition. This work opens the door to a novel metabolic-neuromodulatory avenue that merits confirmation in larger cohorts and establishment of clinical relevance in patients with comorbid neurological and cardiovascular disorders.

## Funding

This work was supported by the Fundamental Research Grant Scheme of the Ministry of Higher Education, Malaysia under the award number FRGS/1/2022/SKK01/UPM/02/4.

## Declaration of Competing Interest

The authors declare no competing interests.

## Data Availability

Deidentified raw datasets of this study are freely available on the Open Science Framework repository at https://doi.org/10.17605/OSF.IO/R97CG.

## Notes

### Competing Interest Statement

The authors have declared no competing interest.

### Clinical Trial

NCT06799260

### Clinical Protocols

https://clinicaltrials.gov/study/NCT06799260

### Author Declarations

Ethical approval was obtained from the local Institutional Review Board of the University Putra Malaysia, approval number JKEUPM-2021-485.

